# Psychological Distress and Metabolomic Markers: A Systematic Review

**DOI:** 10.1101/2022.02.24.22271464

**Authors:** Yiwen Zhu, Shaili C. Jha, Katherine H. Shutta, Tianyi Huang, Raji Balasubramanian, Clary B. Clish, Susan E. Hankinson, Laura D. Kubzansky

**Affiliations:** Department of Epidemiology, Harvard T.H. Chan School of Public Health, Boston, MA, USA; Department of Biostatistics, Harvard T.H. Chan School of Public Health, Boston, MA, USA; Channing Division of Network Medicine, Department of Medicine, Brigham and Women’s Hospital and Harvard Medical School, Boston, MA, USA; Harvard Medical School, Boston, MA, USA; Department of Biostatistics and Epidemiology, University of Massachusetts Amherst, Amherst, MA, USA; Broad Institute of Massachusetts Institute of Technology and Harvard, Cambridge, MA, USA; Department of Social and Behavioral Sciences, Harvard T.H. Chan School of Public Health, Boston, MA, USA

**Keywords:** Psychological distress, anxiety, PTSD, depressive symptoms, metabolomics, systematic review

## Abstract

Psychological distress is a multifactorial construct that refers to non-specific symptoms of depression, anxiety, posttraumatic stress disorder (PTSD), or stress more generally. A systematic review of metabolomic markers associated with distress has the potential to reveal underlying molecular mechanisms linking distress to adverse health outcomes. The current systematic review extends prior reviews of clinical depressive disorders by synthesizing 39 existing studies that examined metabolomic markers for PTSD, anxiety disorders, and subclinical psychological distress in biological specimens. Most studies were based on small sets of pre-selected candidate metabolites, with few metabolites overlapping between studies. Vast heterogeneity was observed in study design and inconsistent patterns of association emerged between distress and metabolites. To gain a more robust understanding of distress and its metabolomic signatures, future research should include 1) large, population-based samples and longitudinal assessments, 2) replication and validation in diverse populations, 3) and agnostic metabolomic strategies profiling hundreds of targeted and nontargeted metabolites. Addressing these research priorities will improve the scope and reproducibility of future metabolomic studies of psychological distress.

**Highlights:**

- Literature on metabolomic markers of distress beyond clinical depression is scarce
- Most existing studies were candidate based and had little overlap of targets
- Vast heterogeneity exists in methods and patterns of findings from studies reviewed
- Critical gaps in sample selection, study design, and methods need to be addressed

## 1. Introduction

### 1.1 Psychological distress

Psychological distress can be characterized by a composite of common psychological conditions, ranging from subclinical symptoms to clinical diagnoses of depression, anxiety, stress, or posttraumatic stress disorder (PTSD). High levels of distress can serve as a marker of impaired mental health or of the presence of common mental disorders, including depression or anxiety. Various forms of psychological distress, defined based on differing diagnoses and measures, are highly comorbid, share similar symptoms, and may be linked to the same underlying pathways of dysregulation (Kalin, 2020); therefore, it is reasonable to hypothesize they might share a similar metabolomic signature.

Various forms of psychological distress have consistently been linked to a range of adverse physical health outcomes, including accelerated aging, cognitive decline, and mortality (Russ et al., 2012; Miller and Sadeh, 2014; Cohen et al., 2015; Roberts et al., 2015, 2017). The most methodologically rigorous work has examined associations of psychological distress with excess risk of cardiometabolic diseases (CMD). Findings have been particularly strong and consistent, with associations evident across animal models, clinical samples, and population-based cohorts (Grippo and Johnson, 2009; Sumner et al., 2015; Golbidi et al., 2015; Levine et al., 2021). Despite this consistency, molecular mechanisms underlying these associations are not well-delineated. One promising hypothesis is that distress may affect downstream metabolism across biochemical domains, (e.g., steroids, amino acids, lipids), leading to dysregulated metabolomic profiles that give rise to excess disease risk (Cohen et al., 2007; Dinoff et al., 2017; Juster et al., 2010).

### 1.2 Metabolomics

Metabolomic markers, or metabolites, refer to a wide range of small molecules that occur in biological fluids and tissues, including peptides, lipids, amino acids, nucleic acids, and many other chemical compounds that can be metabolized (Wishart et al., 2018; Yu et al., 2019). It has been long appreciated that metabolite levels are linked to health and functioning, are influenced by genetics, health, diet, and environmental exposures, and can serve as a signatures of processes that are proximal to disease phenotypes (Gerszten and Wang, 2008; Clish, 2015). In the last 20 years, metabolites have been reliably measured across different biospecimens including plasma, serum, urine, and cerebral spinal fluid (CSF), making it possible to gain greater insight into disease conditions, molecular signatures, and potential therapeutic targets. A metabolomics perspective may also provide important new understanding of key mechanisms and causal pathways underlying the relation between psychological distress and CMD (Humer et al., 2020). Understanding directionality of associations in this context is important as there are likely bidirectional relationships between diseases or psychological conditions and metabolomic profiles.

Several key features of design and analytic approaches distinguish metabolomics studies from one another. First, studies may use a variety of different techniques for metabolomics profiling, such as Nuclear Magnetic Resonance (NMR), and mass spectrometry (MS). Compared to NMR-based methods, MS is thought to more effectively reduce sample complexity and increase detection of metabolite levels especially when a large number of metabolites are assessed simultaneously (Lee et al., 2019; Zhou and Yin, 2016). Second, analogous to other omics studies, both candidate and agnostic approaches have emerged in studies evaluating the relationship between distress and metabolites. A candidate approach involves *a priori* selection of a small to moderate number of metabolites as the focus, and their selection is generally based on specific hypotheses drawn from theory or prior literature. An agnostic approach assesses associations with a large number (typically hundreds) of metabolites simultaneously and detects signals for future follow-up experiments. Third, researchers can use a targeted versus nontargeted approach when measuring metabolites. Most studies to date have utilized targeted strategies, involving quantification of the relative abundance of known metabolites. However, with the recently developed capacity to simultaneously assay hundreds of metabolic markers at once, platforms measuring large-scale metabolomic markers can detect not only known analytes (i.e., a targeted approach), but also unlabeled peaks that can then be mapped to specific metabolites *post hoc*, including structurally novel metabolites (i.e., a nontargeted approach) (Schrimpe-Rutledge et al., 2016). Nontargeted strategies are advantageous especially when the goal of research is to generate new hypotheses. Lastly, while studies may examine the same metabolites, differences in results and interpretations may arise from identifying these metabolites using different types of biospecimen. For example, metabolites concentrations in plasma represent molecules in peripheral circulation whereas metabolites in urine are derived from the degradation and excretion of molecules. Different types of biospecimen may be implicated in different pathways and mechanisms of disease physiology. Without an integrative and standardized pre-processing and analytical procedure, studies looking at different biospecimen types separately may yield seemingly inconsistent results because in fact, they are measuring different pathways (Patti et al., 2012; Zhou et al., 2019).

### 1.3 Associations between psychological distress and metabolomic profiles

Preliminary evidence suggests specific psychological disorders may be linked to unique metabolic signatures in several peripheral tissues. In animal models of depression, tissue-specific biomarkers have been identified; in particular, neurotransmitter and kynurenine metabolite levels in the brain, and amino acid and corticosterone metabolite levels in blood have been linked to depression (Pu et al., 2021). A recent systematic review focused on human studies considering metabolomic markers (from blood, urine, and CSF) in relation to several forms of clinical affective disorders, including major depressive disorder (MDD) and bipolar disorder (BD) (MacDonald et al., 2019). This review included evidence from 266 articles considering 249 metabolites and found 122 metabolites were evaluated in at least two studies. Several key pathways implicated in MDD emerged, including mitochondrial/energy metabolism, neuronal integrity, and signaling/neurotransmission. The review also highlighted a few metabolites showing consistent associations with MDD and BD, such as glutamate and other amino acids, which play key roles in signaling, neurotransmission and energy metabolism. In 2020, a meta-analysis from nine Dutch cohorts (n=10,145 controls and n=5,283 cases with depression) identified 21 metabolites associated with clinically diagnosed depression (Bot et al., 2020).

Findings suggested the metabolomic signature of depression is characterized by lower high-density lipoprotein and higher very-low-density lipoprotein and triglyceride particle levels, highlighting the importance of lipid metabolites in the pathophysiology of depression.

These review articles, both primarily focused on clinical mood disorders, have considerably contributed to our understanding of linkages between psychological distress and metabolomics. However, psychological distress encompasses a broader set of phenotypes beyond mood disorders, including subclinical manifestations of these disorders; our understanding of how psychological distress may influence metabolic processes across this broad spectrum remains limited. To our knowledge, no review to date has considered other forms of psychological distress such as anxiety disorder or PTSD, nor considered subclinical symptoms of these types of distress in population-based samples. It remains unclear whether metabolic markers for clinical depression are similar in other forms of psychological distress or in subclinical distress. In the current systematic review, we address this question by synthesizing existing literature on psychological distress and alterations in metabolomic biomarkers, considering multiple forms of psychological distress at both clinical (e.g., anxiety disorder, post-traumatic stress disorder) and subclinical (e.g., symptoms of anxiety or depression or post-traumatic stress that may not meet criteria for psychopathology) levels. We excluded studies of clinical depression and related disorders given the recently published reviews described above.

### 1.4 Study aims

We aim to summarize and assess literature examining associations between metabolomic profiles in human adults and psychological distress, covering anxiety disorders, PTSD, or subclinical symptoms of depression, anxiety, and PTSD. Objectives include the following: 1) to summarize the aims, study characteristics, and methodological approaches of existing distress and metabolomics studies; 2) to assess converging and diverging evidence from these studies; and 3) to identify critical directions for future work.

## 2. Methods

### 2.1 Eligibility and study selection process

An electronic literature search was performed using PubMed (NLM) and PsychINFO (Ovid) in June 2020. Search terms were selected to identify metabolomics studies of psychological distress, specifically pertaining to subclinical levels of depression as well as clinical or subclinical levels of anxiety and PTSD. All searches were limited to original research conducted in human adults as we were primarily interested in the implications for human diseases.

We conducted a search for studies with subclinical and population-based samples for depression and studies of clinical and population-based samples for anxiety and PTSD (full details on the search strategy are available in the appendix). We excluded two types of studies that were captured by the searches. First, studies measuring metabolite levels using magnetic resonance spectroscopy (MRS) were removed due to general challenges in validating MRS results and varying degrees of reliability depending on the metabolites and brain regions assessed (Dhamala et al., 2019). Second, studies assessing acute stress responses following experimental manipulations were excluded (e.g., alterations in metabolite levels following exposures designed to induce anxiety in patients with phobia or panic disorder), as we were primarily interested in metabolic markers linked with chronic psychological distress.

The Covidence platform was used to organize and perform abstract screening. Two independent reviewers (SJ and RS) reviewed abstracts using standardized criteria. Studies were rejected if they reported findings from animal models, children, or pregnant women. Abstracts that reported findings primarily related to drug effects, herbal treatments, or any sort of intervention and treatment were also excluded. Furthermore, we include only primary research articles, and excluded all commentaries, editorials, opinions, and reviews. We further excluded grey literature, unpublished studies, and articles not available in English, and also studies with a primary focus on mental, physical, or medical conditions other than general psychological distress, subclinical depression, PTSD, and anxiety.

When disagreements regarding a study’s relevance occurred between abstract reviewers, a third rater (LK) evaluated the abstract. Once all abstracts were screened and a set of relevant research articles were collected, both raters performed a full text review applying similar inclusion and exclusion criteria. Finally, new searches and screening were conducted once more before finalizing our analyses to collect and include any additional studies published since initiating our systematic review. **Figure 1** shows the process of study selection.

**Figure 1.**
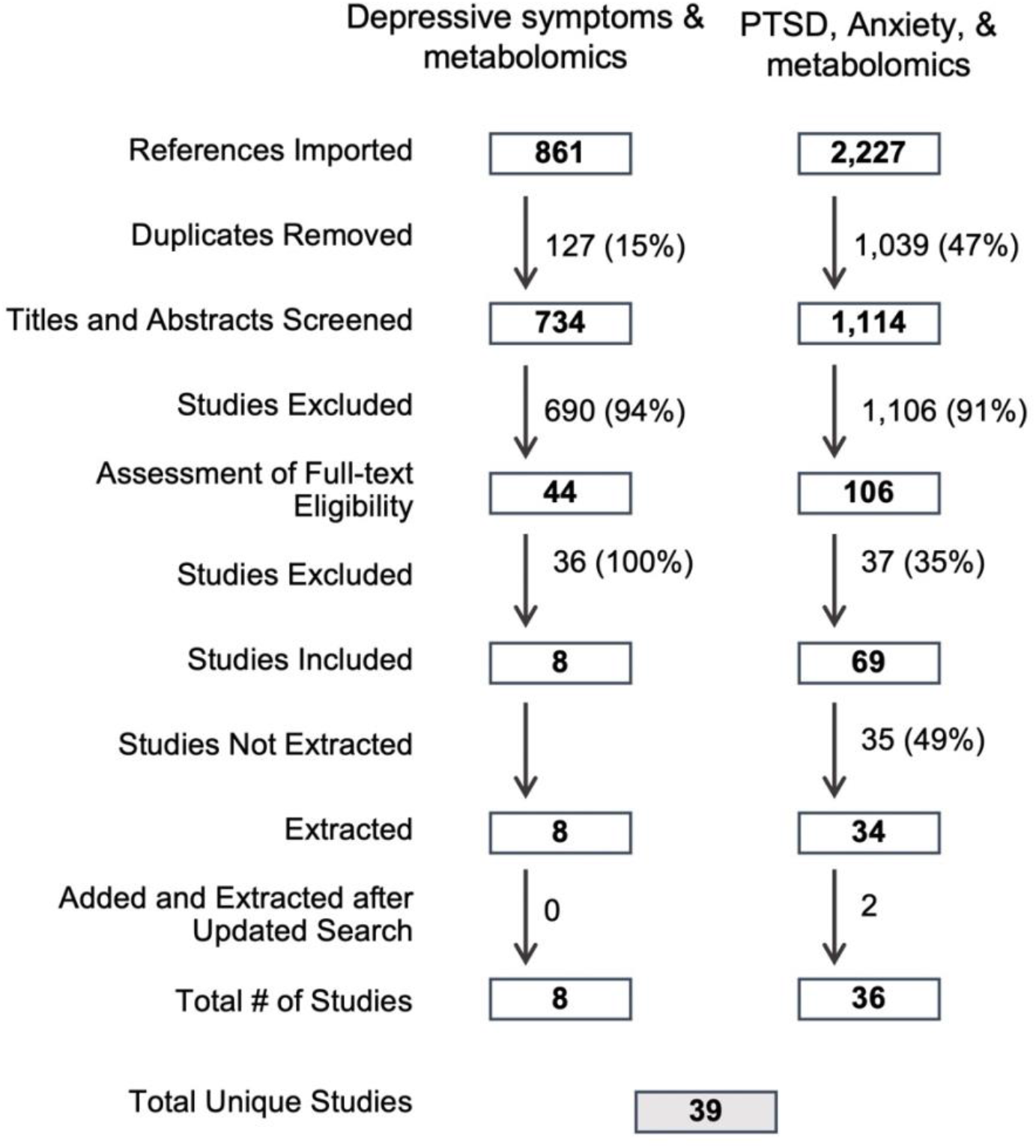
Diagram showing the process of study selection, following the PRISMA guidelines.

### 2.2 Data extraction and analysis

For each study included, we extracted features related to publication details, key population characteristics, study design, measures used to assess distress and metabolomics, statistical analysis, and main findings. Each reported metabolite was entered into the Human Metabolome Database (HMDB, www.hmdb.ca) to extract the corresponding ID, common name, and class. Key characteristics were summarized into separate tables for each of the three distress types: PTSD, anxiety disorders, and subclinical distress including depressive or anxiety symptoms.

Study quality was evaluated using an adapted version of the Newcastle-Ottawa Scale for Cohort Studies (Herzog et al., 2013; Wells et al., 2010) by two independent reviewers (SJ and YZ). The scale was modified to evaluate metabolomic studies in three specific domains: selection, comparability, and outcomes. Specifically, quality of sample selection included the following items: representativeness of study population, sample size, rigor in assessment of exposure, and exclusion of outcome of interest at baseline (when study was longitudinal) or temporality. Study comparability assessment focused on covariate adjustment, i.e., if the most important potential confounders (defined based on prior literature and expertise from our research group, including key determinants of metabolomics and distress – age, sex, and/or race/ethnicity) and additional confounders were considered. Outcome assessment consisted of three items: representativeness of outcome (i.e., scope of metabolite measurement regarding whether candidate vs. agnostic approaches were used), metabolite measurement, and statistical test. Disagreements between raters were resolved by consensus.

## 3. Results

### 3.1 Search results and quality assessments

A database search for studies of subclinical depression and metabolomics identified 861 potentially relevant articles. After excluding 127 duplicate articles, a total of 734 abstracts were screened and 44 articles were further reviewed using the full text. Eight of these met review criteria and were retained for analysis.

A database search for studies of anxiety or PTSD and metabolomics identified 2,227 papers. After removing 1,039 duplicate papers, a total of 1,114 paper abstracts were screened. Of these, 106 studies entered a full-text screening process and a total of 69 studies advanced to the extraction phase. 36 of these met review criteria and were retained for analysis.

Combining results from these two searches yielded a total of 39 unique papers that were included in this review. Of these, 21 primarily targeted PTSD, seven targeted anxiety disorders, and 11 targeted subclinical distress (generally characterized by anxiety or depressive symptoms). If studies included more than one measure of distress, we used the measure which addressed the primary research question. We included fewer studies specifically focused on clinical anxiety disorders compared to studies on clinical PTSD and subclinical symptoms of depression and anxiety, primarily because we excluded studies that experimentally induced short-term responses related to the disorder (such as in phobia or panic disorders). Of the 39 unique reports, two studies utilized an agnostic approach. For these papers, we reported null associations between distress and metabolites only if the same metabolites were also examined in at least two other candidate studies. Given that more than two hundred metabolites were tested in each of the two agnostic studies included in the current review, it was not feasible to examine all tested associations.

Characteristics of all included studies are summarized in **Table 1**. An increasing number of studies were conducted after the year 2000, corresponding to the timeline of technology for assessing metabolites becoming more available and development of various metabolomic platforms (**Figure 2**; Scalbert et al., 2009). Quality assessments completed in the 39 studies revealed nine to be of good quality, 23 to be of average quality, and seven to be of poor quality and at high risk for bias. The full assessment and results can be found in **Table S1**. The least endorsed quality measures were temporality (most studies were cross-sectional) and representativeness of the selected sample (most studies were based in small clinical samples).

**Figure 2.**
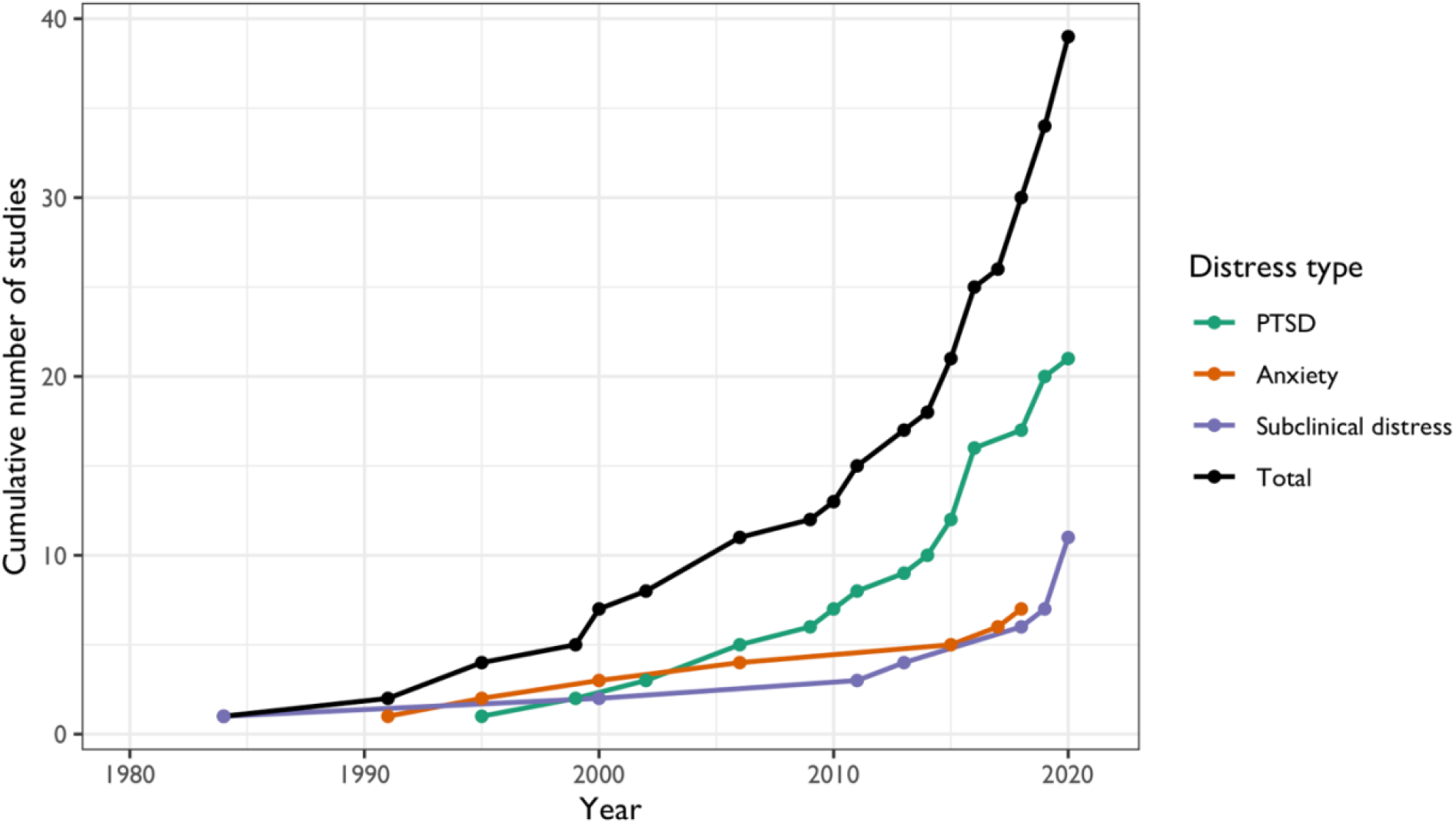
Trend of publication examining psychological distress and metabolic markers.

**Table 1.**
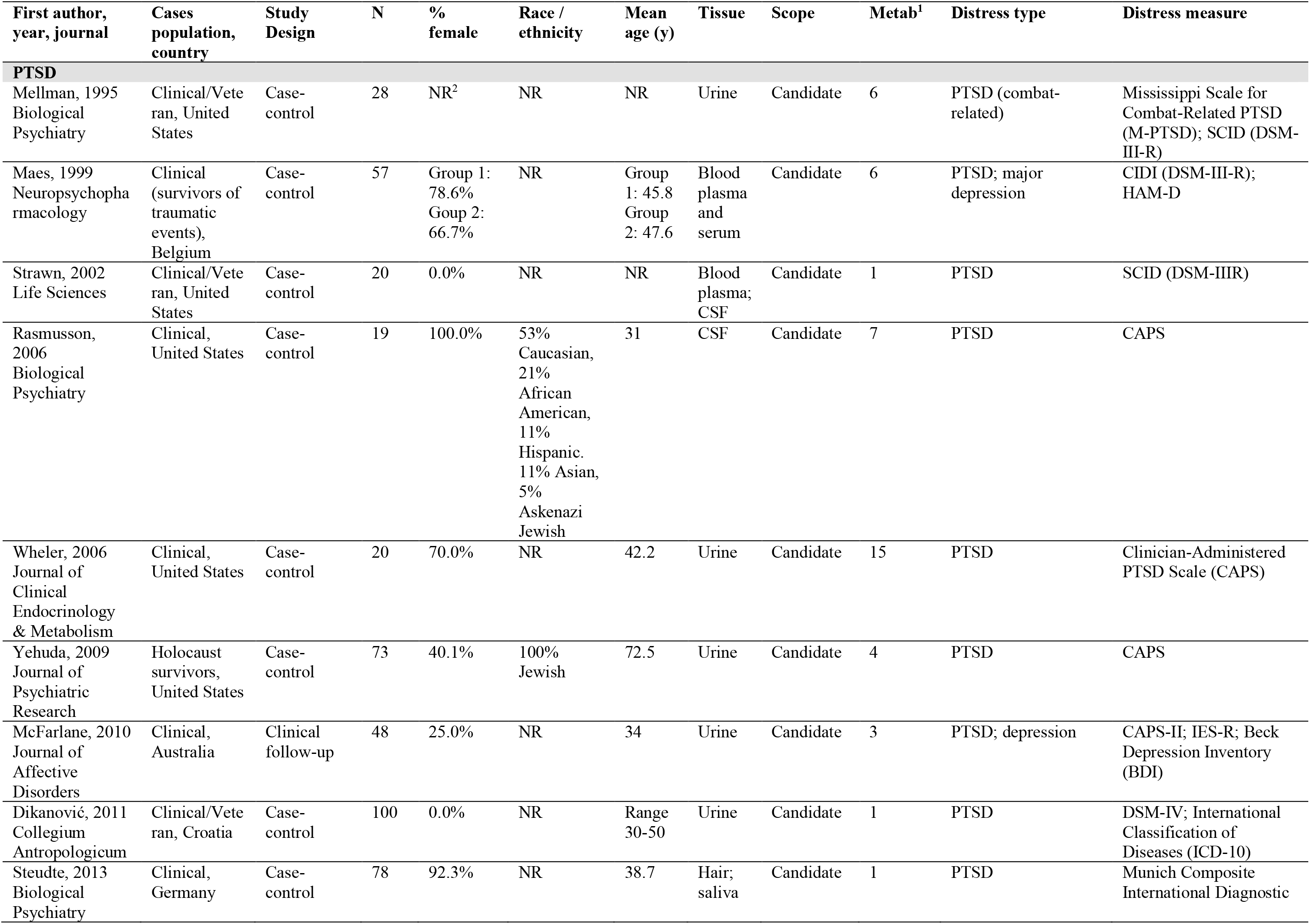
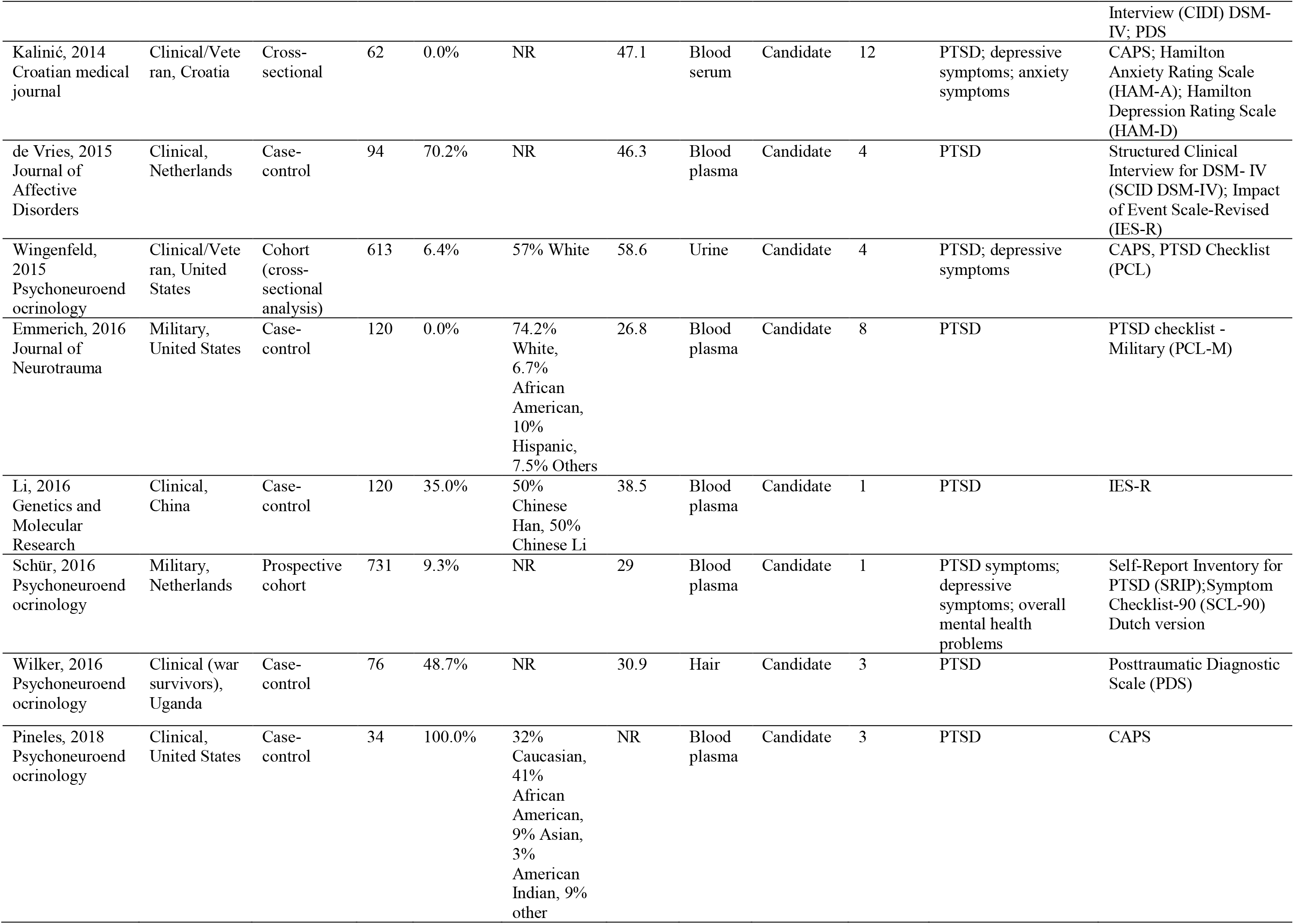
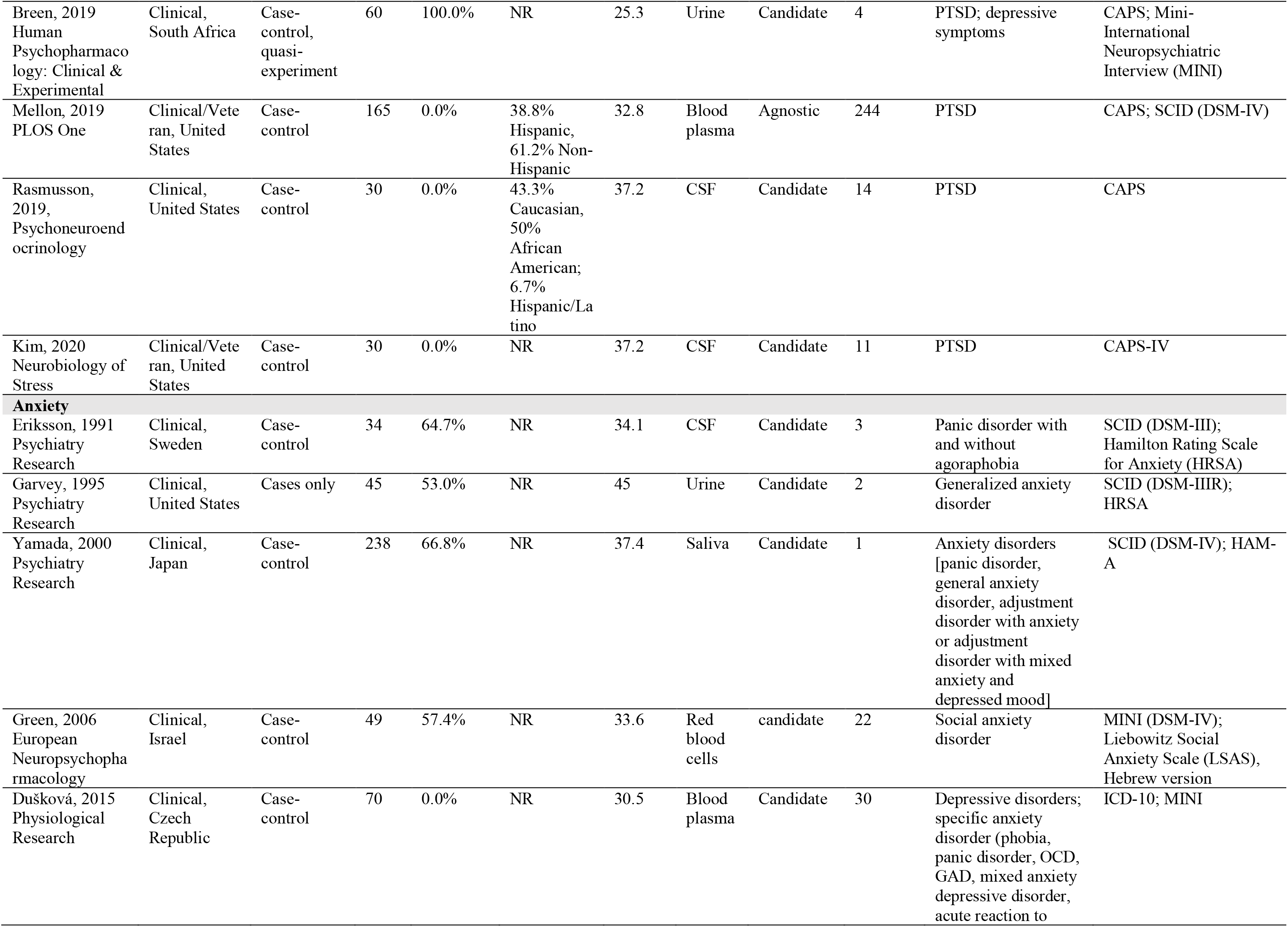
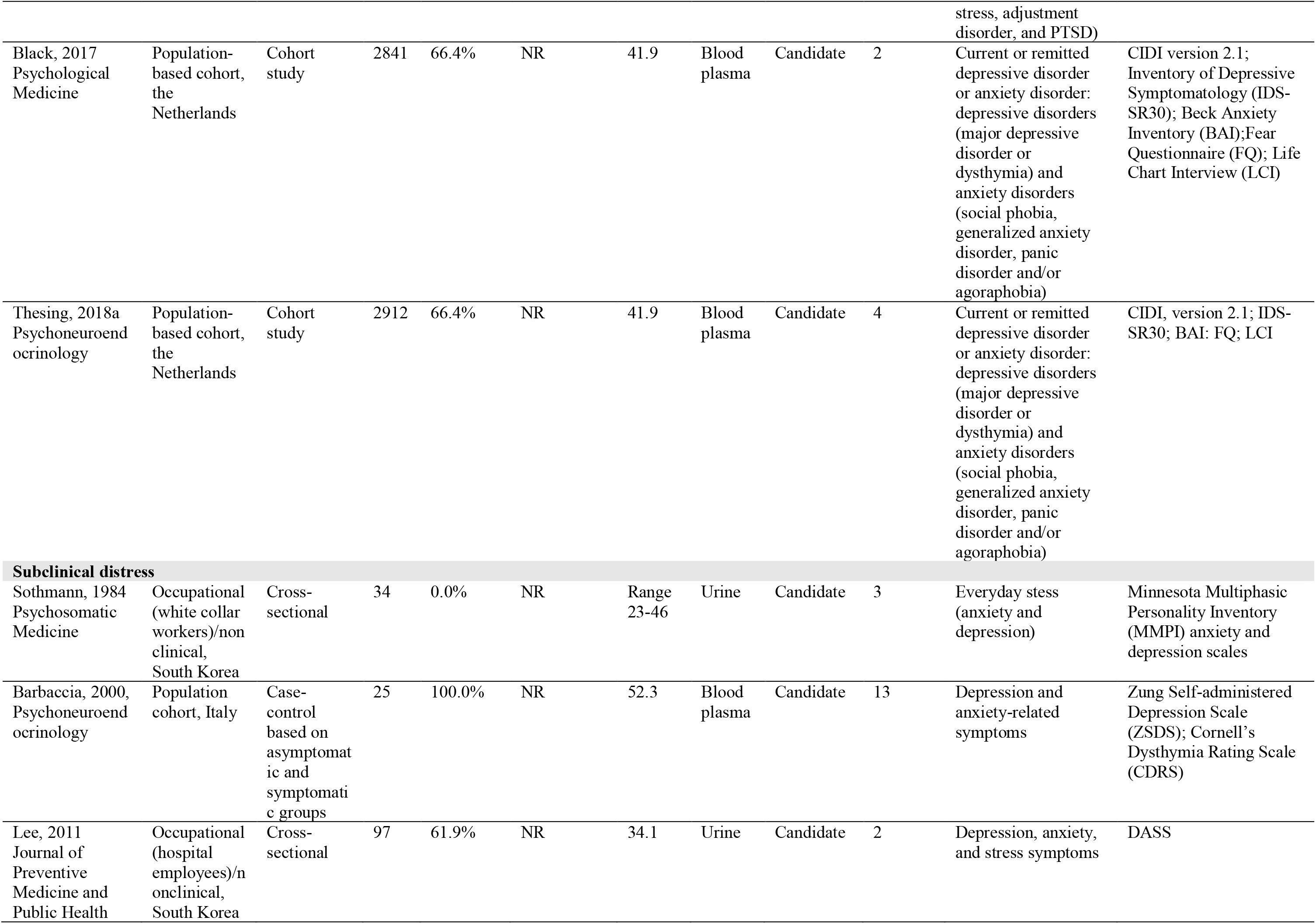
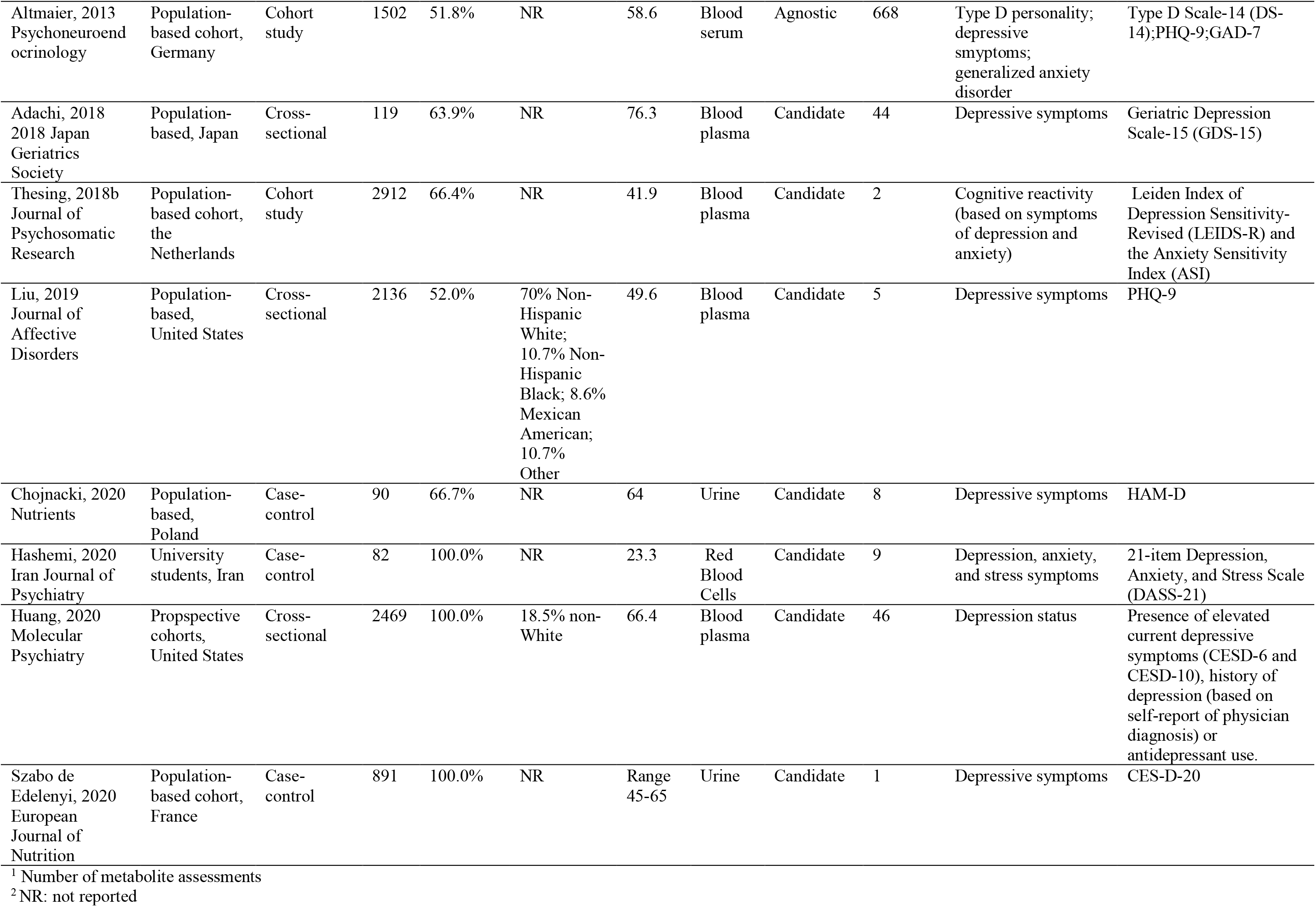
CharacteristisHuangc of studies included in the current review.

Studies that scored low on the quality assessment frequently had inadequate sample sizes and did not adjust for important risk factors. Generally, studies with higher quality scores were those using larger and more representative samples.

### 3.2 PTSD status or symptoms and metabolites

We identified 21 studies. Sample sizes ranged from 19 to 731, but the majority (90%) had fewer than 200 participants. Eleven studies were conducted in civilian clinical populations, while the rest relied on specific populations with elevated levels of trauma exposure and clinical diagnoses of PTSD: nine studies were based in veteran or military samples, and one study included Holocaust survivors. Out of the 17 studies reporting the age distribution of participants, the average age was 39 years. Only one study included a sample of older adults (average age above 60 years; Yehuda et al., 2009). Seven studies included only male, and three studies included only female participants; among studies with both male and female participants, the average percentage of female participants was 44.1% (**Figures S1-S2**).

Most studies used a case-control design (n=17) (**Figure S1C**). Among the remaining four studies, two were analyses of cross-sectional data nested within ongoing cohort studies, one study assessed associations between symptom severity and metabolite levels cross-sectionally in a sample of veterans recruited for that purpose, and one observational study included patients with clinical disorders who were followed and assessed at two additional timepoints after baseline. Metabolites were assayed in a range of biospecimens across the 21 studies: blood plasma (n=8), urine (n=7), CSF (n=4), blood serum (n=2), hair (n=2), and saliva (n=1). Of note, three studies collected more than one type of biospecimen and performed pre-processing and analysis separately for each type (**Figure S3**). Across the 21 studies, only one took an agnostic approach, examining all 244 compounds available on the analytic platform (Mellon et al., 2019). The others evaluated associations with from one to 15 prespecified candidate metabolites.

Measures of PTSD overlapped across a subset of the studies: namely, the Clinician Administrated PTSD scale was used to measure PTSD severity in 11 studies (Blake et al., 1995), and the Structured Clinical Interview for DSM-III-R or DSM-IV was administered in five studies (Spitzer et al., 1992; First and Gibbon, 2004). Other instruments used in more than one study to assess PTSD included the Impact of Event Scale-Revised (Weiss, 2007), PTSD checklist (Blanchard et al., 1996), and the Posttraumatic Diagnostic Scale (Foa et al., 1997). All these measures have been psychometrically validated. While 18 studies primarily focused on the comparison of metabolites between PTSD cases and controls, associations of metabolite levels with PTSD symptom severity on a continuous scale were also examined in 14 studies.

#### 3.2.1 Metabolites in candidate PTSD studies

A total of 57 unique metabolites were examined across the 20 studies of candidate metabolites and PTSD. We were able to match forty-three metabolites to existing HMDB IDs, spanning 10 distinct classes (**Figure 3**). Twelve metabolites or composite measures (e.g., metabolite ratios or aggregate) available in HMDB were selected for study in at least two candidate studies (**Figure 4**). Here, we summarize results regarding the three top metabolite classes with overlap across studies: steroids and steroid derivatives, phenols, and carboxylic acids and derivatives. Metabolite classes that appeared only in one study are not discussed, but a detailed summary of all associations examined is provided in **Table S2**. Direction of associations with PTSD (i.e., when higher PTSD or case status is associated with higher or lower metabolite levels) varies across the different metabolites but are indicated in **Table S2**.

**Figure 3.**
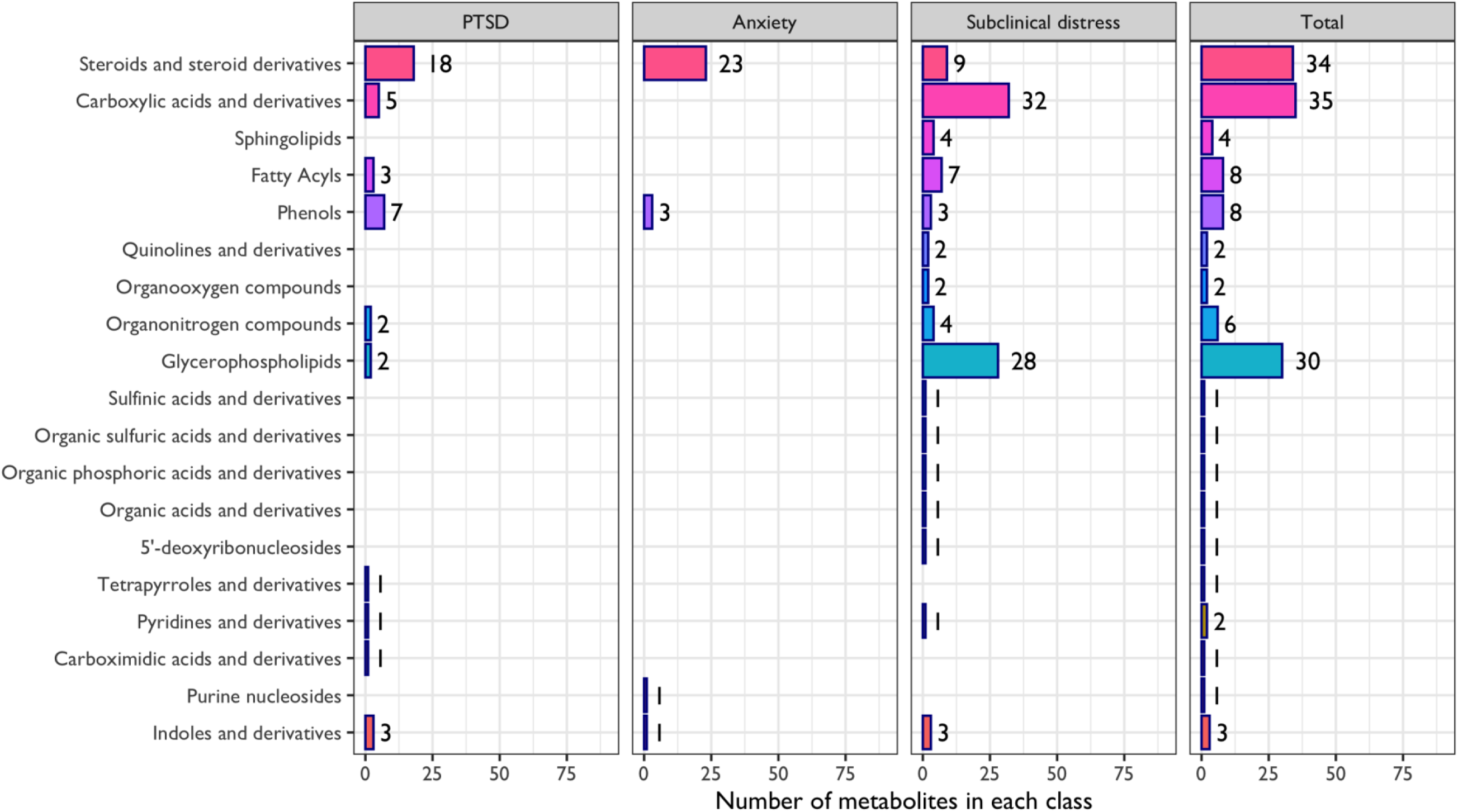
Distribution of HMDB metabolite classes in studies included in the current review.

**Figure 4.**
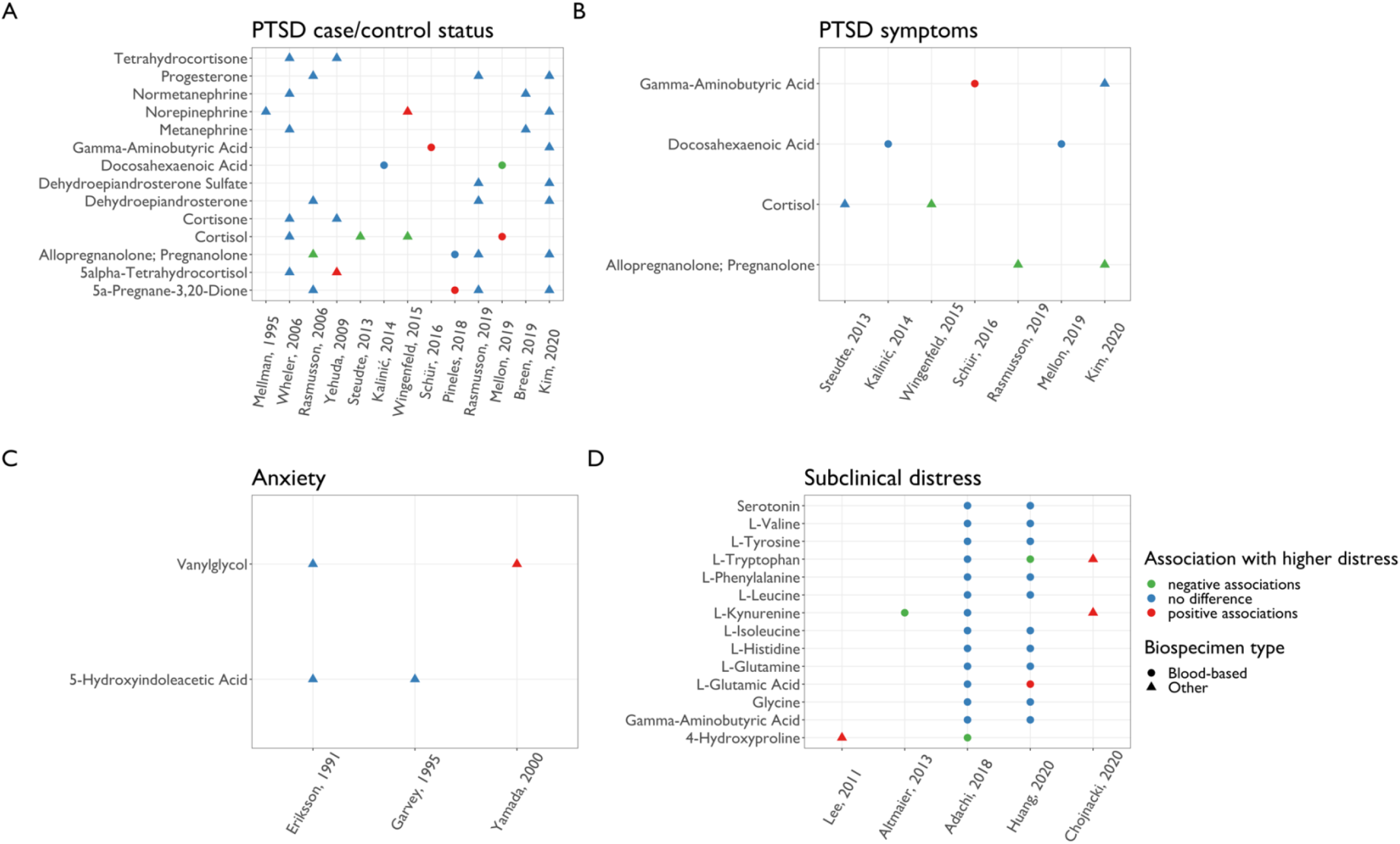
Comparison of associations between psychological distress and metabolites across studies, within each distress type. Metabolites examined in at least two studies within each distress category are shown.

**Figure 5.**
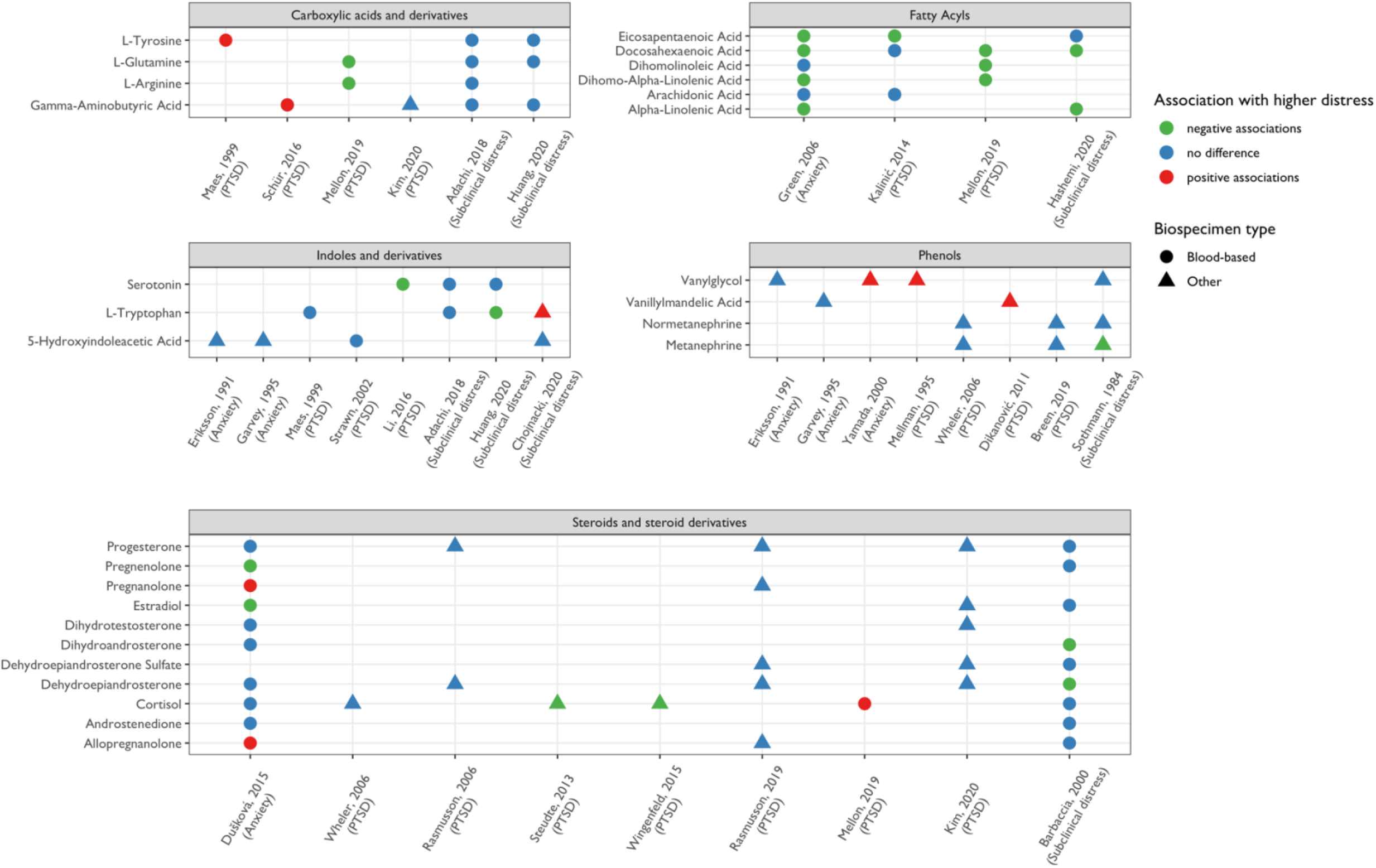
Comparison of associations between psychological distress and metabolites across distress types. Metabolites examined in at least two studies across distress categories are shown.

*Steroids and steroid derivatives.* A total of eight studies examined links between PTSD and 18 steroid metabolites available in HMDB. Four studies tested the hypothesis that deficits in the synthesis of GABAergic neuroactive steroids, such as metabolites of progesterone, may be implicated in the pathophysiology of PTSD (Rasmusson et al., 2006; Pineles et al., 2018; Rasmusson et al., 2019; Kim et al., 2020). Rasmusson et al. (2006) assessed four neurosteroids in CSF (5a-DHP, dehydroepiandrosterone (DHEA), progesterone, Allo+Pregnan), examining differences between nine premenopausal women with chronic PTSD and 10 healthy controls with no trauma history. PTSD patients had lower Allo+Pregnan levels, but no other differences emerged. Another study examined levels of Allo+Pregnan and 5a-DHP in women with PTSD (N=15) compared to trauma-exposed controls (N=19) in blood plasma (Pineles et al., 2018).

Contrary to prior findings in CSF, plasma 5a-DHP levels were higher in PTSD patients but no difference in plasma Allo+Pregnan levels were evident. A follow-up study in 2019 investigated CSF levels of the same neurosteroid metabolites in 13 men with PTSD compared to 17 healthy male controls with trauma history. Although no between-group differences were identified, an inverse correlation between Allo + Pregnan levels and a continuous measure of PTSD symptom severity emerged (Rasmusson et al., 2019). Lastly, Kim et al. (2020) identified a negative correlation between CSF Allo+Pregnan levels and PTSD symptom severity among trauma-exposed men with and without PTSD (N=30) but found no difference in 5a-DHP levels or DHEA. Three of these four studies also assessed associations between metabolite ratios and PTSD. The Allo+Pregnan/5a-DHP ratio was inversely associated with PTSD in two studies (Rasmusson et al., 2006; Pineles et al., 2018) and not associated in one study (Rasmusson et al., 2019). The Allo+Pregnan/DHEA ratio was also inversely associated with PTSD in both studies that included this measure (Rasmusson et al., 2006, 2019).

Investigators have also considered metabolites related to glucocorticoid metabolism. Four studies examined the associations between specific glucocorticoid metabolites and PTSD status or symptoms (Wheler et al., 2006; Yehuda et al., 2009; Steudte et al., 2013; Wingenfeld et al., 2015). Overall, results were mixed: Wheler et al. (2006) found no difference in urinary levels of 14 cortisol metabolites between 10 PTSD patients and 10 controls matched by age and gender. In a sample of aging Holocaust survivors (N=51) evaluating urinary levels of four glucocorticoid metabolites, Yehuda et al. (2009) identified one positive association between PTSD symptom severity and 5a-THF. Wingenfeld et al. (2015) analyzed urinary cortisol levels in a large cohort recruited at two Veterans Affairs medical centers (N=613) found cortisol levels were *lower* in PTSD patients. A fourth study evaluating hair and salivary cortisol also found *lower* hair cortisol levels among those with PTSD in comparison to non-trauma exposed controls, but no association with salivary measures (Steudte et al., 2013).

*Phenols or catecholamine metabolites.* Six studies examined levels of seven phenols available in HMDB. Of three studies assessing norepinephrine levels in CSF or urine samples (Kim et al., 2020; Mellman et al., 1995; Wingenfeld et al., 2015), only one found a positive association with PTSD status which suggested norepinephrine levels in urine may be higher in PTSD patients; however, this study did not find associations with either epinephrine or dopamine (Wingenfeld et al, 2015). Two studies using small samples (including 10-20 individuals with PTSD and approximately the same number of healthy controls) examined metanephrine and normetanephrine levels from urine and found no link to PTSD status (Wheler et al. 2006; Breen et al. 2019). Notably, while Mellman et al. (1995) did not identify differences in nocturnal or 14- hour excretion levels of urinary norepinephrine or MHPG between PTSD patients and controls (N=28), their analyses did find PTSD patients had a larger difference between their nocturnal and daytime MHPG levels, reflecting potential dysregulation during sleep. One study measured VMA, an epinephrine and norepinephrine metabolite; findings from 50 patients with chronic PTSD and 50 healthy controls showed that VMA derived from urine was higher in patients (Dikanović et al., 2011).

*Carboxylic acids and derivatives.* Five studies assessed a total of five carboxylic acids and derivatives. The only metabolite analyzed in at least two studies was gamma-aminobutyric acid (GABA). While Kim et al. (2020) found GABA levels in CSF did not differ between trauma-exposed men with and without PTSD in a small sample (N=30), Schür et al. (2016) identified a positive association between GABA levels in plasma and PTSD symptoms in a large sample of military personnel (N=731).

#### 3.2.2 Agnostic analyses

Out of the 21 studies on PTSD, only one study performed an agnostic analysis of metabolomic profiles (Mellon et al., 2019). The authors identified 244 compounds in plasma available in both the discovery (N=103) and test samples (N=62) and compared group-level differences between combat trauma-exposed male veterans with and without PTSD, matched on age. Identified markers were primarily related to glycolysis and fatty acids uptake and metabolism. Among 33 identified markers, only cortisol and Docosahexaenoic acid (DHA) were also examined in the candidate studies of PTSD described above, with cortisol levels being higher among PTSD patients in the discovery group but not the test group, and DHA being lower among PTSD patients in both groups. Additionally, two metabolites, cortisone and dehydroepiandrosterone sulfate (DHEA-S), showed consistently null relationships with PTSD in this agnostic analysis as well as in other candidate studies (Wheler et al., 2006; Yehuda et al., 2009; Rasmusson et al., 2019; Kim et al., 2020).

### 3.3 Anxiety disorders and metabolites

A total of seven studies documented the relationship between anxiety disorders and metabolites in clinical populations. Studies were performed in six different countries with sample sizes ranging from 34 to 2,912 individuals. The majority of studies (57%) were small, with less than 100 participants. Notable exceptions were two large population-based cohorts that included 2,841 and 2,912 individuals recruited from the Netherlands Study of Depression and Anxiety (NESDA) (Black et al., 2017; Thesing et al., 2018a). All studies were conducted among individuals in early to mid-adulthood, with mean age across samples ranging from 30.5 to 45 years. One study consisted of only male participants. In studies with both male and female participants, the average percentage of female participants was approximately 62.5% (**Table 1**).

Four studies defined “anxiety disorders” as having a clinical diagnosis of any anxiety disorders including social phobia, generalized anxiety disorder (GAD), panic disorder and/or agoraphobia (Black et al., 2017; Dušková et al., 2015; Thesing et al., 2018a; Yamada et al., 2000). One study focused specifically on panic disorder (Eriksson et al., 1991), one on social anxiety disorder (Green et al., 2006), and one on GAD (Garvey, 1995). Diagnoses were made using the Structured Clinical Interview (for DSM-II, DSM-IIIR, or DSM-IV), the Composite International Diagnostic Interview (CIDI), or the International Classification of Diseases, Tenth Revision (ICD-10). Anxiety symptoms were most often evaluated using the Hamilton Rating Scale for Anxiety (HRSA) or the Beck Anxiety Inventory (BAI).

No agnostic studies of anxiety disorders and metabolites were conducted. Three studies used blood plasma, and one study each used urine, saliva, red blood cells, and CSF to measure metabolite concentrations (**Table S3**).

#### 3.3.1 Metabolites in candidate studies

A total of 48 unique metabolites were identified in the seven studies. Of these, 37 matched existing HMDB IDs. These 37 metabolites belong to five different metabolite classes. Only two of these were documented in at least two studies: Vanylglycol (MHPG) belonging to the phenols metabolite class, and 5-Hydroxyindoleacetic acid belonging to the indoles and derivative class (**Figure 4C**).

MHPG was assessed in both saliva and CSF. Yamada et al. (2000) assessed salivary levels of MHPG in patients with diagnosed anxiety disorders (including panic disorder, GAD, adjustment disorder with anxiety or adjustment disorder with mixed anxiety and depressed mood) and found higher salivary MHPG concentrations in patients versus controls. In contrast, Eriksson et al. (1991) found no significant differences in CSF MHPG levels in patients with panic disorder compared to controls.

5-Hydroxyindoleacetic acid (5-HIAA) was assessed in CSF (Eriksson et al., 1991) and urine (Garvey, 1995). Neither study revealed significant relationships of 5-HIAA concentrations with panic disorder or with total symptom scores.

### 3.4 Subclinical distress and metabolites

A total of eleven studies documented the relationship between depressive or anxiety symptoms and metabolites (**Table 1**). All studies were conducted in sub-clinical or population-based cohorts, with three comprising occupational samples and one comprising students. Studies were performed in nine different countries with sample sizes ranging from 25 to 2,912 individuals. More than a third of these comprised large population-based cohorts with over 1,000 participants. Mean age across studies ranged from 23.3 years to 76.3 years, with most studies performed in middle age and older participants. Four study samples included only female, and one study included only male participants; among studies with both males and females, the average percentage of female participants was approximately 60.5%.

Distress symptoms were assessed using a variety of different measurement tools. The 21- item Depression, Anxiety, and Stress Scale (DASS-21) was administered in two different studies (Hashemi et al., 2020; Lee et al., 2011). Depression screeners such as the CES-D (CES-D-20, CES-D-10, or CES-D-6), and PHQ-9 were also used in two or more studies (Altmaier et al., 2013; Huang et al., 2020; Liu et al., 2019; Szabo de Edelenyi et al., 2020). Altmaier et al. also utilized Type D personality, a measure of general psychological distress characterized by both social inhibition and negative affectivity.

Ten of the eleven studies (91%) were candidate studies, pre-selecting study metabolites based on previous research. The remaining study (Altmaier et al., 2013) performed an agnostic assessment of Type D personality using 668 metabolites. Metabolites were assayed from blood plasma (N=5), urine (N=4), blood serum (N=1), or red blood cells (N=1) (**Table S4**).

#### 3.4.1 Metabolites in candidate studies

A total of 106 unique metabolites were identified in the 11 studies pertaining to subclinical distress. Most metabolites (N=98) were matched to existing HMDB IDs and classified into 14 different metabolite classes. Fourteen of these were selected as candidates in at least two studies, with eleven belonging to the carboxylic acids and derivatives metabolite class (**Figure 4**). The remaining metabolites belonged to the following classes: indoles and derivatives, fatty acyls, steroids and steroid derivatives, and organooxygen compounds classes.

##### Carboxylic acids and derivatives

The relationship of **carboxylic acids and derivative metabolites** with subclinical distress was examined in three studies. Of the 11 metabolites examined, only hydroxyproline and glutamic acid showed significant trends with distress in at least one study.

Hydroxyproline was evaluated as a candidate metabolite in two studies (Adachi et al., 2019; Lee et al., 2011). Adachi et al. (2019) categorized a sample of Japanese older community dwelling adults into two groups experiencing high vs. low depressive symptoms as measured by the Geriatric Depression Scale-15, then examined the association with levels of plasma amino acid-related metabolites, including hydroxyproline. Lee et al. (2011) examined the relationship between urinary hydroxyproline and depression, anxiety, and stress symptoms, as measured by the Depression Anxiety Stress Scale (DASS), in hospital employees from South Korea. Both studies found significant associations, albeit in biospecimen-specific and in opposite directions: Adachi et al. showed a negative relationship between plasma hydroxyproline levels and depressive symptoms (Adachi et al., 2019) while Lee et al. found urinary hydroxyproline concentrations were higher among those with high levels of stress symptoms but not with either depressive or anxiety symptoms (Lee et al., 2011).

Glutamic acid was considered as a candidate metabolite in two studies as well (Adachi et al., 2019; Huang et al., 2020), but findings were inconsistent. Huang and colleagues (2020) assessed the association of depression status (yes/no), measured by reporting either elevated current depressive symptoms, history of depression (based on self-report of physician diagnosis), or antidepressant use, with glutamic acid (as well as other candidate metabolites from plasma) in three independent samples of postmenopausal women in the US. While this study found average levels of plasma glutamic acid were higher in the depressed cases relative to controls, the Adachi et al. (2019) study described above observed no significant differences between those with and without depressive symptoms. Six additional carboxylic acids and derivative metabolites were also assessed in these studies, but none were significantly associated with any measure of depression in either study.

##### Indoles and derivative metabolites

Two **indoles and derivative metabolites**, tryptophan and serotonin, were examined in two or more studies (Adachi et al., 2019; Chojnacki et al., 2020; Huang et al., 2020). Details of Adachi et al. (2019) and Huang et al. (2020) are described above. Chojnacki and colleagues (2020) used the Hamilton Depression Rating Scale (HAM-D) to characterize depressive symptom levels and evaluated urinary metabolite levels in middle-aged adult without mood disorders, older aged adults without mood disorders, and older adults with mild and moderate depressive symptoms.

While all three studies evaluated tryptophan, associations varied across all reports. One found significantly lower levels of plasma tryptophan in the depressed cases relative to controls (Huang et al., 2020), another reported higher urinary levels of tryptophan in older adults with depressed mood compared to middle-aged adults without depression symptoms (Chojnacki et al., 2020), and the third study found no associations between plasma tryptophan and depressive symptoms among older adults (Adachi et al., 2019). Two studies considered serotonin as a candidate metabolite but found no significant relationships with depressive symptoms (Adachi et al., 2019; Huang et al., 2020) .

##### Other metabolites

DHA and kynurenine, belonging to the **fatty acyls and organooxygen compounds classes,** respectively, were also assessed in two or more studies and association were generally inconsistent across studies. DHA levels were evaluated in two studies (Hashemi et al., 2020; Thesing et al., 2018b). Hashemi et al. (2020) assessed the relationship of DHA in red blood cells with self-reported stress and anxiety symptom levels, as measured by the DASS, in Iranian university students, comparing a group with elevated subclinical stress and anxiety to matched controls with low levels of depression, stress, or anxiety symptoms. Thesing et al. (2018b) investigated the association of depression and anxiety sensitivity, as measured using the Leiden Index of Depression Sensitivity-Revised (LEIDS-R) and the Anxiety Sensitivity Index (ASI), with plasma DHA in a large cohort of healthy adults. The first study found lower DHA levels among participants with elevated subclinical stress and anxiety, while the second study found no evidence of an association.

Kynurenine levels were assessed in two studies (Adachi et al., 2019; Chojnacki et al., 2020) and associations with depressive symptoms were similarly inconsistent. Urinary concentrations of kynurenine were higher among elderly participants with elevated depressive symptoms when compared to non-depressed younger adults (Chojnacki et al., 2020), but plasma concentrations were not associated with depressive symptom severity in the second candidate study (Adachi et al., 2019).

#### 3.4.2 Agnostic analysis

Altmaier et al. (2013) performed an agnostic analysis using metabolomic panels by Metabolon, Inc. and Biocrates to identify a signature for Type D personality in a population-based cohort of 1502 German adults. Using an agnostic assessment of the associations of 668 serum metabolites, the study found significantly lower levels of kynurenine in individuals with versus without Type D personality. Moreover, when looking at the two subscales of Type-D personality, there was a positive association between cortisol and social inhibition and a negative association between cortisol and negative affectivity.

### 3.5 Concordance of Metabolite Associations Across Distress Phenotypes

Across studies of PTSD, anxiety disorders, and subclinical distress, 28 metabolites were examined in studies considering relationships with at least two different forms of distress. These 28 metabolites span five metabolite classes: steroids and steroid derivatives, fatty acyls, carboxylic acids and derivatives, phenols, and indoles and derivatives. Out of the 28 metabolites, 21 were reported to be significantly associated with a form of distress in at least one study.

#### 3.5.1 Steroids and steroid derivatives

Across nine studies, eleven steroids and steroid derivatives were assessed in relation to the different forms of distress. Cortisol was the most frequently studied metabolite (included in a total of six studies) but findings were inconsistent. For example, while cortisol was negatively correlated with PTSD in two candidate studies using hair and urine samples, respectively (Steudte et al., 2013; Wingenfeld et al., 2015), higher serum cortisol was associated with lower negative affectivity and higher social inhibition in a large agnostic study (Altmaier et al., 2013); another agnostic study also found higher plasma cortisol levels among individuals with PTSD.

Adding to the inconsistencies, in a small candidate study found no association of subclinical distress with plasma cortisol levels (Barbaccia et al., 2000).

Progesterone and dehydroepiandrosterone (DHEA) were also commonly studied, examined in five studies. None found an association of progesterone with any form of distress, regardless of study design, biospecimen type, or analytic platform. Apart from one small study (N=25) documenting a negative association for depression and anxiety symptoms with DHEA, no other studies found evidence of associations. Related, DHEA-sulfate was assessed in three studies, and associations with any form of distress were consistently null. Other metabolites in this class were either included in two or fewer studies or had mixed results across studies.

#### 3.5.2 Fatty acyls

Six fatty acyl metabolites were studied across four studies, with two focusing on PTSD, one on anxiety disorder, and one on subclinical distress. While associations between distress and fatty acyls varied across studies examining the same distress type (i.e., PTSD), some consistency was observed across different forms of distress. For example, six of the omega-3 fatty acids examined in relation to social anxiety disorder by Green et al. (2006) were also analyzed in other studies, and five showed consistent directions of associations in at least one other study. While some studies failed to identify an association, no conflicting directions of associations occurred for any of the fatty acyl metabolites.

#### 3.5.3. Carboxylic acids and derivatives

L-Tyrosine, L-Glutamine, L-Arginine, and GABA were examined in relation to various distress types in a total of six studies. While the two studies on subclinical distress (Adachi et al., 2019; Huang et al., 2020) did not find a significant relationship, single studies on PTSD identified significant associations without replication. GABA, the most frequently studied metabolite in this class, was examined in four studies, with two examining associations with PTSD and two examining subclinical distress. Out of these, only one identified a significant association with PTSD (Schür et al., 2016). Additionally, L-Glutamine and L-Arginine levels were lower among individuals with PTSD (Mellon et al., 2019) but none of these metabolites were significantly associated with subclinical distress levels in other studies.

#### 3.5.4 Phenols

Four phenol metabolites, all of which pertain to catecholamine metabolism, were assessed across eight studies of different distress types. Two out of four studies examining levels of MHPG found positive associations. Specifically, Mellman et al. (1995) found significant daytime to nocturnal differences in metabolites levels between PTSD patients and controls. One study of anxiety found higher MHPG concentrations in patients versus controls (Yamada et al., 2000). However, other studies evaluating this metabolite in relation to anxiety or subclinical distress found no significant concentration differences between groups (Sothmann and Ismail, 1984; Eriksson et al., 1991). Normetanephrine was assessed in three studies, all of which found no association with distress. Out of the three studies that assessed levels of metanephrine, only one identified a negative association (Sothmann and Ismail, 1984); the two studies that included VMA also yielded inconsistent results (Garvey, 1995; Dikanović et al., 2011).

#### 3.5.6 Indoles and derivatives

Three indoles and derivatives (5-HIAA, serotonin, and L-tryptophan) were examined in eight studies considering various forms of distress. All four studies including 5-HIAA found no association with any forms of distress, while results concerning relationships with serotonin and L-tryptophan were mixed. Serotonin levels were lower among individuals with PTSD in one study (Li et al., 2016) but no associations were evident in two large studies of subclinical depression. Conflicting findings regarding L-tryptophan emerged from the two studies on subclinical distress: while Huang et al. found lower levels of plasma L-tryptophan associated with higher distress, another study found higher levels of urinary L-tryptophan associated with higher levels of distress (Chojnacki et al., 2020).

## 4. Discussion

Data extracted from the 39 studies provide the current state of evidence for associations between psychological distress and various metabolic markers. Our synthesis did not yield a clear and robust set of metabolites reliably associated with one or more forms of psychological distress. However, failure to find a common set of metabolites altered in relation to various forms of distress is likely due, at least in part, to the widely varying methodological approaches, biospecimens, and metabolites selected for assessment across the studies that have been done to date. Specifically, few candidate studies assessed the same metabolites and even the overlap of metabolite classes across studies was limited. Additionally, most studies were heterogeneous with regard to their underlying population, design, biospecimen type, laboratory procedures, and data processing pipelines; moreover, many studies were small, such that even when the same metabolites were examined, possibilities for replication and validation were limited. Our findings strongly suggest the need for more unified and systematic examination of these relationships.

That said, while the heterogeneity across studies suggests we cannot yet draw any conclusions about a common set of distress-linked metabolites, a substantial amount of information can be drawn from existing studies to inform future investigations.

Summarizing across metabolite classes, we identified several notable patterns. First, although many studies examined steroids and steroid derivatives, regardless of the type of distress considered or the specific metabolites examined, most associations were null. Even investigations of metabolites linked to well-documented pathways underlying stress responses, such as cortisol regulation, yielded mixed results. Second, similar inconsistencies across findings emerged for several other metabolite classes across the various forms of distress, including catecholamine metabolites (e.g., MHPG), serotonin and its metabolites, and amino acids. Third, when comparing across distress types, there was suggestive evidence of consistent associations between fatty acids and several forms of distress. In the following sections, we unpack each finding in more detail and discuss recommendations for future studies.

### 4.1 Metabolite findings within and across distress types

Comparing studies looking at similar forms of distress, we largely observed discordant results for most candidate metabolites evaluated. For example, across studies of PTSD and anxiety disorders, the most commonly examined metabolite class was steroids and their derivatives; however, we did not see consistent patterns of associations even when comparing only among studies of anxiety or only among studies of PTSD that looked at the same metabolites. Of note, CSF levels of a composite measure, Allo+Pregnan, were negatively associated with PTSD severity in two studies among men and women, although no association was evident with PTSD case-control status in the same two studies. Somewhat surprisingly, cortisol-related metabolites, long identified as part of a stress-linked pathway, were not consistently associated with PTSD status or symptoms, nor with anxiety as examined in one study. Furthermore, across studies, few individual steroid or their derivative metabolites were consistently associated with any forms of distress. Beyond potential differences in design or analytic approach (see below for more detailed discussion), one potential explanation is that cortisol metabolites specifically may not be independently associated with measures of distress, but dysregulation of the entire pathway could play a key role in stress physiology, as shown in a differential network analysis from relevant recent studies that explored this approach (Shutta et al., 2021). Similarly, although catecholamines play a central role in sympathetic nervous system activation and are heavily implicated in stress response regulation, we failed to see any robust links among the studies reviewed here. Moreover, while numerous studies examining subclinical distress evaluated associations with candidate carboxylic acids and derivatives, few meaningful or consistent associations were found across studies.

Compared to the body of work linking metabolic markers with clinical depression which has identified key pathways related to neurotransmission or energy metabolism (MacDonald et al., 2019), studies of subclinical distress are somewhat sparse and current evidence for consistent associations is limited. Most studies considered either subclinical depression or subclinical symptoms of depression and anxiety. Among studies of subclinical depression, at least 10 metabolites selected as candidates by two studies (Adachi et al., 2019; Huang et al., 2020) failed to demonstrate significant associations. In contrast, the previous systematic review of clinical depression found significant associations with eight of these metabolites in two or more studies (MacDonald et al., 2019). One possible explanation for differences in findings across these different ways of characterizing distress is that metabolite differences evident in individuals experiencing severe and clinical levels of depression may not be as potent or detectable among individuals with subclinical depression. However, another possibility is that detection rate of metabolites could also vary by analytical platforms and biospecimen. Because most studies use different platforms and many use different biospecimen, efforts to compare findings across studies and forms of distress are further complicated. It is also possible that measures of subclinical versus clinical distress capture different underlying constructs depending on what features of the disorder are measured. In that case, biological signatures of these phenotypes may not completely overlap, analogous to differences that have been observed between the genetic architecture of minimally phenotyped versus clinically defined depression (Cai et al., 2020).

Several additional issues may be at play regarding the divergence in findings regarding metabolite association with clinical versus subclinical depression. Lack of consistency could be attributed to the narrow scope of metabolic function captured by studies of candidate metabolites; as the field moves forward with agnostic approaches based in larger samples, additional insights regarding metabolic differences in pre-clinical and clinical populations are likely to emerge. Another issue could be differences in sample sizes and statistical power: large studies (i.e., n>1,500) considering subclinical depression with either tryptophan (Huang et al., 2020) or kynurenine (Altmaier et al., 2013) each produced results consistent with those observed in clinical studies of depression (MacDonald et al., 2019). Furthermore, in a recent discovery and validation study that was performed across large subclinical datasets (Shutta et al., 2021), relationships between depressive status and GABA and serotonin were identified, consistent with findings in studies of clinical depression.

Other factors could also introduce confounding and noise into metabolomic analyses of psychological distress, rendering it more difficult to find consistency across studies. As noted by prior reviews (Davison et al., 2018; MacDonald et al., 2019), a key factor is whether studies take account of psychotropic medication status. Medications may affect associations of interest because their therapeutic effects could induce changes in metabolomic profiles. Of the 39 studies included in this review, twelve did not provide specific information about medications that may impact distress and metabolomics associations (i.e., psychotropic medications). Nineteen studies either excluded all individuals taking psychotropic medication or required a washout period (i.e., asked participants to stop taking their medications) prior to sample collection. The remaining eight studies included individuals using psychotropic medications. Of these, two PTSD case-control studies of veterans’ cohorts consisted entirely of patients using psychotropic medication. The remaining six studies focused on antidepressant use and considered the variable as a confounder or evaluated its effects in secondary and sensitivity analyses. While these reports generally revealed no significant impact of antidepressant use (Black et al., 2017; Mellon et al., 2019; Wingenfeld et al., 2015), one of the larger studies did find that in a model simultaneously adjusting for three different depression indicators, associations with amino acids were strongest with antidepressant use compared to depressive symptoms (Huang et al. 2020). In general, 49% of studies in this review included currently unmedicated individuals. However, among studies that did include individuals who are medicated, information on medication was not consistently available, not consistently assessed, and/or not consistently included in analyses. As a result, our ability to determine how psychotropic medication use impacts observed associations of psychological distress with metabolite alterations is limited. Moreover, psychotropic medication use varies widely across different forms of distress, often depending on severity and type of distress. Of note, MacDonald et al. (2019) found antidepressant medication status did not significantly impact findings of altered metabolite levels related to MDD or BD.

Although overlap in findings of metabolite alterations across distress types was limited, we observed several intriguing patterns that may be potential targets for future follow-up studies. First, across the four studies that assessed fatty acyls, a generally consistent picture emerged whereby higher levels of fatty acyls were mostly associated with lower psychological distress. Of note, all four studies analyzed blood samples from a clinical population. This finding suggests not only that reliable metabolite alterations may occur but also that lipids may be an important component of a molecular signature of psychological distress. This inference is supported by other work linking aberrations in lipid profiles to clinical depression (Dinoff et al., 2017; Bot et al., 2020) as well as to higher risk of cardiovascular diseases (Laaksonen et al., 2016; Xu et al., 2016). Taken together, these findings may suggest that lipids are involved in the underlying pathophysiology linking elevated CMD risk with distress. Nonetheless, because only four studies examined fatty acyls and each metabolite was included in at most three studies, more rigorously designed follow-up studies are needed to further validate this finding. Second, for some metabolite classes such as carboxylic acids and indoles, results were largely divergent across distress types. For example, differences in findings about GABA or 5-HIAA across distress types could be attributed to both heterogeneity across studies and potential distress-specific signals.

### 4.2 Study designs and epidemiologic characteristics

As noted throughout, the body of work to date is characterized by great heterogeneity in study designs and epidemiological characteristics. Such differences can make any direct comparisons difficult or unwise. Thus, a key finding from this review is the critical need to consider these factors carefully in future studies. Here, we discuss several key aspects of design, including sample size, tissue type, selection of metabolites, and measurement of distress. Of note, in the current review, studies often differ on many of these elements, making it difficult to pinpoint a specific reason for any heterogeneity in findings.

First, limited sample sizes and consequently insufficient power likely contributed to the failure to detect associations in some studies. In brief, among the reviewed studies, those with larger versus smaller samples were more likely to find positive associations of various metabolite levels with distress. For example, a large sample of middle-aged postmenopausal women found higher depressive symptoms levels were associated with higher plasma glutamate levels (Huang et al., 2020) but this association was not evident in a sample of 152 older community dwelling adults (Adachi et al., 2019). On the other end of the spectrum, underpowered studies are also more likely to yield more biased estimates and potentially exaggerated effects (Gelman and Carlin, 2014). Given most studies in the current review were relatively small, an important focus future work will be to implement metabolomic assessments in large, population-based cohorts.

A second source of heterogeneity relates to variation in tissue type used across studies. For example, a number of studies found urinary tryptophan and kynurenine were higher in individuals with higher depressive symptoms (Chojnacki et al., 2020) but associations were not evident in studies measuring these metabolites in plasma (Adachi et al., 2019). Use of different tissue types may also help explain inconsistent findings regarding associations of cortisol and its metabolites with distress. Measures of metabolites from different biospecimens could reflect different metabolic processes, such as those occurring with acute responses versus chronic dysregulation, or in central versus peripheral processes. Furthermore, concentration of amino acids in CSF or plasma may be influenced by a variety of activities including diet and metabolic processes that control the absorption, transport, degradation and excretion of metabolites, whereas concentrations in urine are largely influenced by the rate of excretion of these molecules (Fonteh et al., 2007). Thus, if alterations in metabolite levels occur only after chronic exposure and reflect long-term functional changes, it might be difficult to observe associations in studies considering only acute responses to distress or using different biospecimens. Related, even if the same types of biospecimens were analyzed, measurements of basal differences may be less reflective of some relative dysregulation compared to change over time, such as the nocturnal- daytime differences of MHPG levels assessed in a study of PTSD (Mellman et al., 1995). Thus, careful attention is needed regarding which biospecimens and measures of metabolites are used.

To mitigate variability arising from these differences, studies should also consider matching factors such as fasting status, date, and time of blood draw.

A third source of heterogeneity is the use of different metabolomic platforms, a problem present in the field of metabolomics as a whole. For example, in the 47 cohorts represented in the COnsortium of METabolomics Studies (COMETS; Yu et al., 2019), the world’s largest metabolomics consortium, at least 15 different analytic platforms were used to collect metabolomics data. Different platforms do not typically evaluate the same set of metabolites. In fact, when comparing the three most commonly used platforms, Yu et al. (2019) found only modest overlap in the metabolites measured across them, with only 14 metabolites measured by all three. In the set of six studies collecting large-scale metabolomics data that were included in the current review, we found four different platforms were used; given limited overlap of metabolites evaluated across platforms, comparability across studies is constrained. There is a pressing need for research that addresses comparability across metabolomics platforms to evaluate more precisely similarities and differences in whether and how various forms of distress may be associated with metabolic alterations.

A fourth source of heterogeneity is how distress is measured, including the use of instruments designed to capture clinical vs non-clinical levels of distress and the assessment of formal diagnoses of disorders versus the use of symptom-level gradients. For example, while prior studies reported associations between PTSD symptom severity and the combined measure of Allo+Pregnan, no such associations were evident in studies comparing PTSD cases and controls. Additionally, for PTSD research specifically, the selection of controls remains an important question. By definition, PTSD occurs only in the context of experienced trauma. Thus, some studies defined the control group as individuals who experienced trauma but not PTSD, while other studies compared individuals meeting clinical criteria for having PTSD with a control group of individuals with no trauma history. Determining which control group is most appropriate would involve developing an understanding of whether trauma itself, even in the absence of any additional psychological distress, might be associated with certain metabolic alterations, and whether researchers are primarily interested in dysregulation that occurs in the context of trauma.

A fifth source of heterogeneity is that existing candidate studies use an inconsistent approach to metabolite selection. While most studies provided justification for why they considered certain sets of metabolites, they did not discuss why these metabolites might be more pertinent to the underlying question of interest than other metabolites. The lack of overlap between studies overall also suggested that researchers are generally more incentivized to pursue novel hypotheses, instead of conducting replication or validation studies. In the omics era, reproducibility of findings is increasingly important. An additional challenge with assessing reproducibility is the requirement that validation or replication studies consider the variability of metabolite concentrations due to both technical measurement error and changes in lifestyle and environmental factors, while using similar study populations and pursuing similar analytic approaches (Perng and Aslibekyan, 2020). That said, ultimately it will be important to consider associations across highly diverse populations.

Lastly, differences in analytic method and covariate adjustment may also lead to different degrees of bias and interpretations of study results. For example, in the agnostic metabolomic study of PTSD, Mellon and colleagues (2019) adjusted for a range of potential confounders, including medication use, comorbid depression (to account for the independent effects of PTSD), and physiological markers. However, other candidate studies that examined the same metabolites generally did not adjust for any additional lifestyle or physiological factors. Among the 39 studies included in the current review, most adjusted for age, sex, or race/ethnicity, while only half additionally accounted for biobehavioral factors (e.g., physical activity, diet). Ten studies included no covariates, merely reporting between-group differences. Such variation makes it challenging to compare estimates across studies. Thus, we must interpret some results as only crude observed differences and can consider only a more limited set of studies as a source of unbiased estimates of the associations of interest.

### 4.4 Recommendations for future studies

Findings from this systematic review highlight the need for large, systematic studies of psychological distress and metabolomics that use a consistent set of methods and platforms.More specifically, we make three key methodological recommendations for future studies. First, diverse study samples and large, population-based cohorts should be prioritized. While studies in our review represent samples from 18 different countries, many are quite small and 74% do not report information regarding race or ethnicity. Consequently, considering consistency of findings across diverse populations is not currently possible. Moreover, among even the large population cohorts to date that do provide racial and ethnic information, all include a majority of non-Hispanic white participants. Recruiting diverse and representative populations can enhance our understanding of natural variations in metabolite levels and pathways related to psychological health (Reavis et al., 2021). Studies of preclinical distress have utilized large cohorts such as the Nurses’ Health Study II and Women’s Health Initiative (Huang et al., 2020), NESDA (Thesing et al., 2018a), and the National Health and Nutrition Examination Survey (Liu et al., 2019), but community-based cohorts addressing clinical PTSD and anxiety disorders are lacking. To date, studies of PTSD and metabolomics have generally been conducted in small clinical samples, and studies of anxiety disorder have not yet been conducted with samples of older adults.

Second, regarding the measurement of metabolomics, we recommend performing agnostic analyses based on large-scale platforms that can provide broader coverage of the metabolome. Such analyses allow for simultaneous measurement of hundreds of metabolites in plasma or other tissue types. We found only two studies that have utilized this high-throughput approach to date. With the introduction of international collaborations such as COMETS (Yu et al., 2019), datasets can be aggregated to produce well-powered, large-scale studies linking metabolomics profiles not only with disease endpoints but also with social and psychological factors. Such datasets can also be used to examine metabolite levels across various tissue types and platforms to standardize and optimize collection methods. In addition to expanding the scope of metabolomic platforms, we recommend implementing studies that not only identify relationships (i.e., discovery) but also validate these relationships using rigorous criteria to demonstrate that associations can be replicated. Our recent study provides an example of this; a metabolome-wide agnostic approach was used to assess metabolomic profiles associated with psychological distress using a discovery and validation design, identifying eleven metabolites with validated associations with psychological distress (Shutta et al., 2021). Additionally, given high variability in metabolite concentrations due to both technical variations and fluctuations in external environments, future studies should carefully assess measurement error and provide coefficients of variation in reporting.

Third, well-powered longitudinal analyses are needed to address causal and mechanistic questions. Specifically, future studies should evaluate the chronicity of psychological distress and assess whether higher levels of distress are associated with change in metabolites over time as well as with the persistence of metabolomic changes over time. Such studies should carefully consider sources of potential confounding, may also consider life course and whether these changes are evident only at particular developmental periods (e.g., among older adults), and assess if such changes may be modified or “reversed” if distress is appropriately treated or remits on its own.

### 4.5 Limitations

There are several limitations to note for this review. First, due to the heterogeneity across studies with regard to methods and platforms and biospecimens, we could not conduct a quantitative synthesis. Second, because most studies excluded individuals who were taking distress-relevant medications, we were unable to assess the role of pharmacological treatment in the dysregulation of metabolite levels. Lastly, the scope of this review is constrained by the criteria used for study selection. As with most systematic reviews, this limitation impacts the generalizability of our findings to other populations, such as children and individuals with clinical depression.

### 4.6 Conclusion

Findings from our systematic review highlight the potential of and need for examining metabolite profiles linked to psychological distress beyond clinical depression. Through summarizing characteristics, methods, and results of 39 existing studies, the review points to a number of important future directions that will make it possible to conduct a more unified, systematic analysis of distress metabolomics. Adequately powered population-based longitudinal studies with multidimensional measures of distress and large-scale assessments of metabolomics are needed to validate existing findings, resolve inconsistencies, and generate novel hypotheses for future research.

## Data Availability

All data produced in the present work are contained in the manuscript.

## Acknowledgements

We thank Rocky E Stroud (Department of Epidemiology, Harvard T.H. Chan School of Public Health) for his contribution to this project, especially throughout the early stages of article screening and data extraction.

