## Supplementary material for "Psychological Distress and Metabolomic Markers: A Systematic Review": 20220224_Supplement_Distress-Metabolomics-Systematic-Review.docx

**Supplemental Materials**

**Search strategy**

The search terms for chronic distress:

((Depressi*[tiab] OR Depression [Mesh] OR Depressive Disorder[Mesh] OR depressive symptoms[tiab] OR depressed [tiab] OR psychological distress[tiab] OR psychological stress[tiab] OR Stress, Psychological[Mesh] OR chronic distress[tiab] OR chronic stress[tiab] OR negative emotion*[tiab] OR Emotional Adjustment[Mesh] OR Ptsd[tiab] OR Psychological Trauma[Mesh] OR Emotions[Mesh] OR Anxiety[Mesh] OR Anxiety[tiab] OR negative mental state[tiab] OR negative affect[tiab] OR Loss of Emotional Control[tiab])

AND (Metabolom* [tiab] OR plasma metabolite [tiab] OR lipid metabolism [tiab] OR Metabolite[tiab] OR metabolic pathway[tiab] OR endogenous metabolite[tiab] OR mass spectrometry[tiab] OR Gas-chromatography [tiab] OR Liquid-chromatography [tiab] OR NMR[tiab] OR GC-MS [tiab] OR LC-MS [tiab] OR "Metabolic Networks and Pathways"[Mesh] OR nuclear magnetic resonance[tiab])

AND (Humans[Mesh] OR men [tiab] OR women [tiab] OR adults [tiab] OR people [tiab])

AND (Population-based[tiab] OR subclinical[tiab] OR general population [tiab] OR cohort [tiab] OR non clinical [tiab] OR healthy population [tiab] OR community [tiab] OR controls [tiab]))

The search terms for anxiety and PTSD:

("Stress Disorders, Post-Traumatic/epidemiology"[Mesh] OR "Stress Disorders, Post-Traumatic/psychology"[Mesh] OR "Psychological Trauma"[Mesh] OR “Ptsd”[tiab] OR post traumatic stress disorder [tiab] or posttraumatic stress disorder [tiab] or post-traumatic stress disorder [tiab] or psychological trauma [tiab] or traumatic experience [tiab] or anxiety [tiab] or anxious[tiab] OR internalizing [tiab] OR anxiety disorder[tiab])

AND (Metabolom* [tiab] OR plasma metabolite [tiab] OR lipid metabolism [tiab] OR Metabolite[tiab] OR metabolic pathway[tiab] OR endogenous metabolite[tiab] OR mass spectrometry[tiab] OR Gas-chromatography [tiab] OR Liquid-chromatography [tiab] OR NMR[tiab] OR GC-MS [tiab] OR LC-MS [tiab] OR "Metabolic Networks and Pathways"[Mesh] OR nuclear magnetic resonance[tiab])

AND (‘Humans’[Mesh] AND ‘Adult’ [Mesh])

**Quality assessment questions**

*Items with an asterisk next to it correspond to a score of 1. See **SuppTable 1** for a complete scoring rubric and scores for each study included in the current review.

Selection

1) Representativeness of the exposed cohort
   a) truly representative of the average _______________ (describe) in the community*
   b) somewhat representative of the average ______________ in the community *
   c) selected group of users e.g., nurses, volunteers
   d) no description of the derivation of the cohort

**Scored 1 if the selection of the cohort was truly or somewhat representative of the average population; scored 0 if the exposed cohort was selected from a specific group such as nurses, physicians, etc., or no description was provided*

2) Sample size
   a) adequately powered to detect a difference *
   b) underpowered or no justification provided

**Scored 1 if the sample size was justified and satisfactory based on adequate power (80% or above); scored 0 if the sample size was not satisfactory or justified based on power < 80%. Power calculations were performed using sample sizes given in each study and corrected for the number of metabolites tested, holding effect size to be OR=1.32, which was the median effect estimate reported in a large empirical study assessing depression and metabolomics (Shutta et al., 2021).*

3) Ascertainment of exposure
   a) secure record (e.g, surgical records) *
   b) structured/clinician interview*
   c) written self-report
   d) no description

**Scored 1 if exposure (psychological distress variables) were ascertained using secure records or structured/clinical interviews; scored 0 if exposure (psychological distress variables) were ascertained using self-report, or no description was provided*

4) Demonstration that outcome of interest was not present at start of study
   a) yes*
   b) no

**Scored 1 if the study excluded participants with psychological distress at baseline; scored 0 if participants with psychological distress were not excluded at baseline*

Comparability

5) Comparability of comparison groups on the basis of the design or analysis
   a) study controls for age; sex; or race/ethnicity the most important factor *
   b) study controls for any additional factor (physical activity; smoking; diet; medication; alcohol use.)*
   c) not used or no description provided

**Scored 1 if the study adjusted for age; sex; race/ethnicity; scored 2 if the study adjusted for any additional factor such as physical activity; smoking; diet; medication; alcohol use; scored 0 if the study did not consider other factors*

Outcomes

6) Representativeness of the measured outcome
   a) large, agnostic, diverse set**
   b) selective, candidate, literature-based set *
   c) self-selected, without justification
   d) no description

**Scored 2 if outcome metabolites were selected from a large agnostic set; scored 1 if outcome metabolites were selected from a literature-based candidate set; scored 0 if the outcome metabolites were self-selected without justification, or description was provided*

7) Was the outcome measured well?
   a) yes, measured well and validated with a CV *
   b) no

**Scored 1 if outcome metabolites were measured well and validated; scored 0 if validation was not completed, or no description was provided*

8) Statistical test
   a) The statistical test used to analyze the data is clearly described and appropriate, and the measurement of the association is present, including confidence intervals and the probability level (p-value)*, including corrections for multiple comparisons
   b) The statistical test used to analyze the data is clearly described and appropriate, and the measurement of the association is present, including confidence intervals and the probability level (p-value)*, but missing multiple testing corrections
   c) The statistical test is not appropriate or incomplete.
   d) The statistical test is not described

**Scored 1 if the statistical test used to analyze the data is clearly described and appropriate, and the measurement of the association is present, including confidence intervals and the probability level (p-value); scored 0 if the statistical test used to analyze the data was no appropriate or complete, or no description was provided*

**Supplemental Tables**

*Due to the size of the tables, we have included them as excel files.*

Supplemental Table 1. Assessment of study quality based on the Newcastle-Ottawa scale

Supplemental Table 2. Metabolite results in 21 studies of PTSD

Supplemental Table 3. Metabolite results in 7 studies of anxiety disorders

Supplemental Table 4. Metabolite results in 11 studies of subclinical distress

**Supplemental Figures**

Figure S1. Characteristics of studies included in the systematic review. A) Study populations; B) Countries where the studies were performed, grouped by continent; C) Study sample designs.

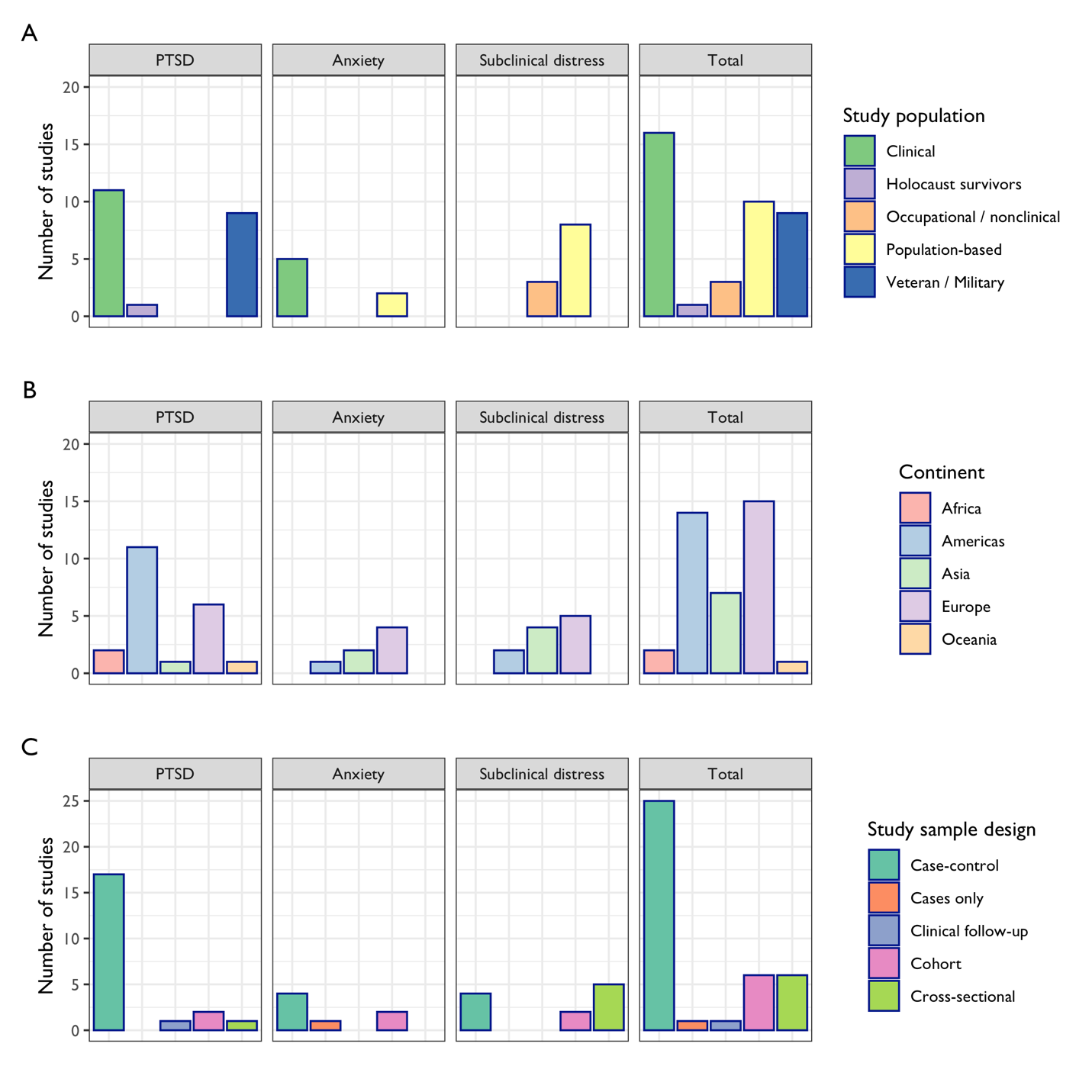

*Note.* Information provided in Panel C correspond to the design of study sample selection instead of the analyses. Cross-sectional studies refer to studies that did not involve separate recruitment for cases and healthy controls and were not embedded in any population-based cohorts. Quantitative traits were measured and examined in the entire study sample.

Figure S2. Characteristics of study samples included in the systematic review. A) Sample sizes; B) Percentage of female participants; C) Mean age of participants. If a range was provided, the mid-point of the range was reported.

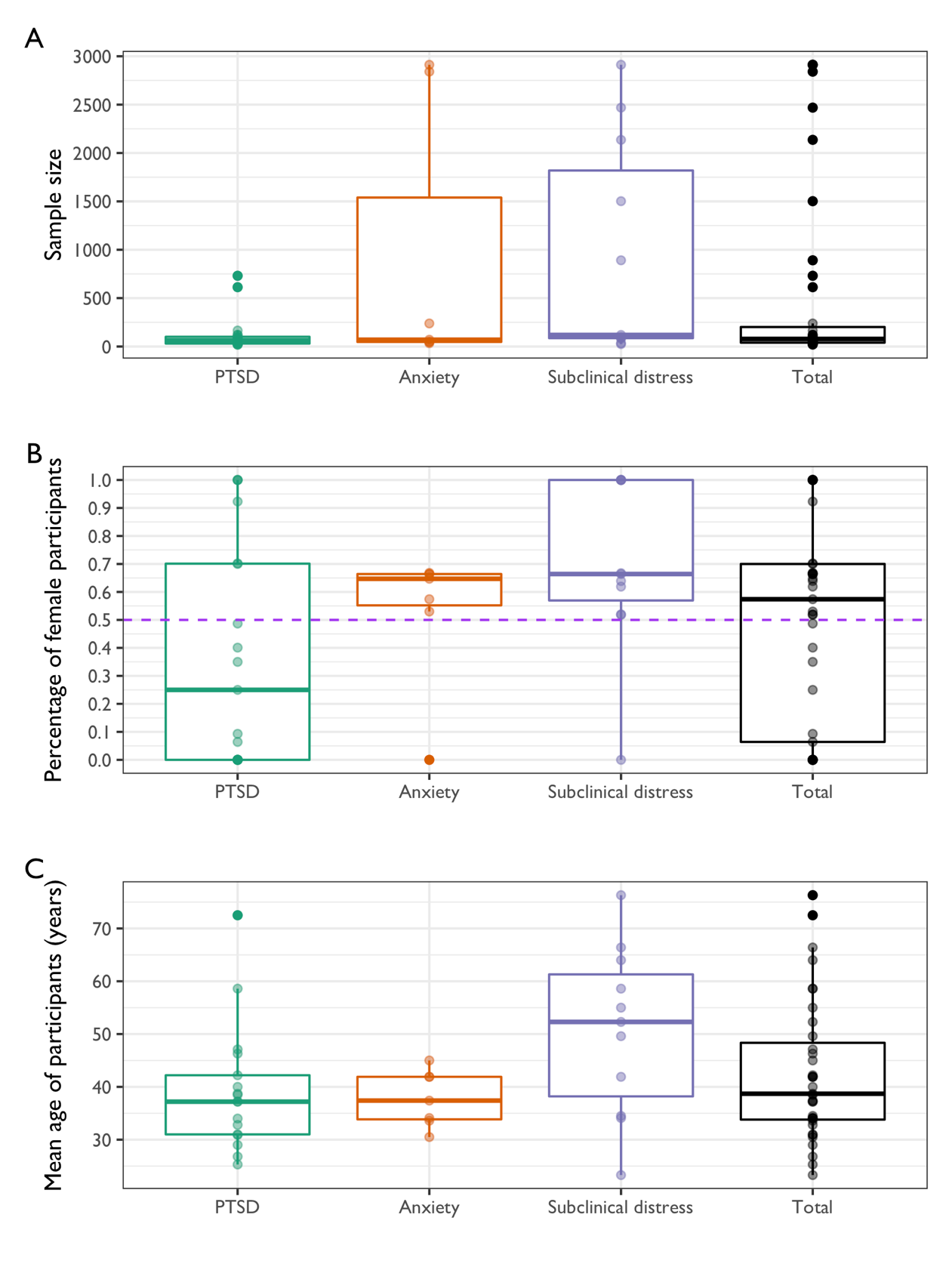

Figure S3. Distribution of biospecimen types analyzed in the included studies.

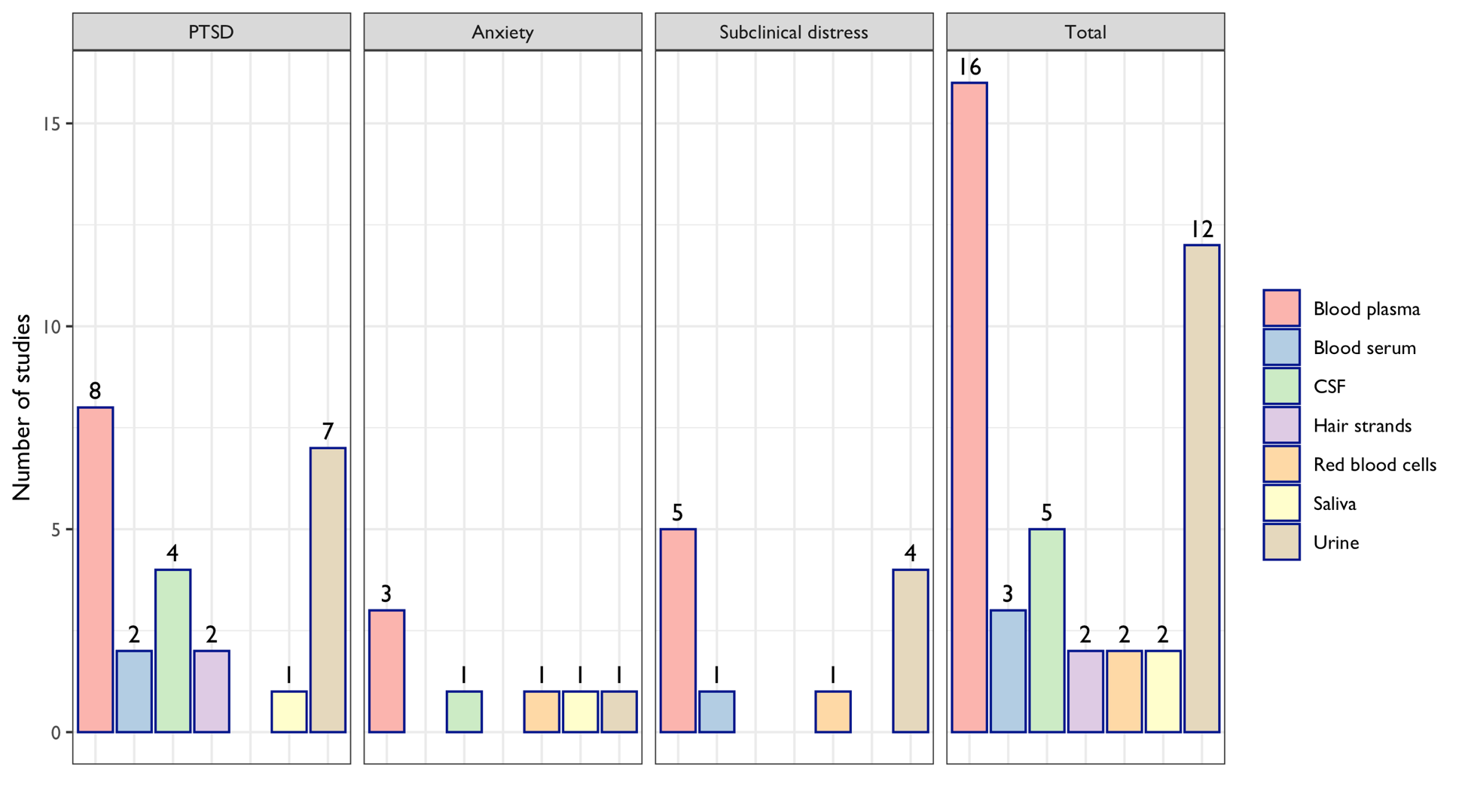
